# Mapping brain endophenotypes associated with idiopathic pulmonary fibrosis genetic risk

**DOI:** 10.1101/2022.03.25.22272932

**Authors:** Ali-Reza Mohammadi-Nejad, Richard J. Allen, Luke M. Kraven, Olivia C. Leavy, R. Gisli Jenkins, Louise V. Wain, Dorothee P. Auer, Stamatios N. Sotiropoulos

## Abstract

**Background:** Idiopathic pulmonary fibrosis (IPF) is a serious disease of the lung parenchyma. It has a known polygenetic risk, with at least seventeen regions of the genome implicated to date. Growing evidence suggests linked multimorbidity of IPF with neurodegenerative or affective disorders. However, no study so far has explicitly explored links between IPF, associated genetic risk profiles, and specific brain features.

**Methods:** We exploited imaging and genetic data from more than 32,000 participants available through the UK Biobank population-level resource to explore links between IPF genetic risk and imaging-derived brain endophenotypes. We performed a brain-wide imaging-genetics association study between the 17 known loci of IPF genetic risk and 1,248 multi-modal imaging-derived features, which characterise brain structure and function.

**Findings:** We identified strong associations between cortical thickness and white matter microstructure and IPF risk loci in chromosomes 17 (*17q21*.*31*) and 8 (*DEPTOR*). Through co-localisation analysis, we confirmed that the associated neuroimaging features and IPF share a single causal variant at the chromosome 8 locus. Exploratory post-hoc analysis suggested that forced vital capacity may act as a partial mediator in the association between the *DEPTOR* variant and white matter microstructure, but not between the *DEPTOR* risk variant and cortical thinning in the anterior cingulate.

**Interpretation:** Our results reveal for the first time associations between IPF genetic risk and differences in brain structure, for both cortex and white matter. Differences in tissue-specific imaging signatures suggest distinct underlying mechanisms with focal cortical thinning in regions with known high *DEPTOR* expression unrelated to lung function, and more widespread microstructural white matter changes consistent with hypoxia or neuroinflammation with potential mediation by lung function.

**Funding:** This study was supported by the NIHR-funded Nottingham Biomedical Research Centre and the UK Medical Research Council.

Research in context

Evidence before this study
Idiopathic pulmonary fibrosis (IPF) is a condition in which the lungs become scarred (fibrosed). Although IPF primarily affects the lungs, co-occurrence of impaired brain function, such as cognitive decline, increased risk of neurodegenerative disorders, cerebrovascular accidents, depression, and anxiety have been reported. The nature of this association is unclear and no post-mortem or in-vivo investigations have been performed to explore this directly. At the genetic level, 17 regions of the genome have been found to drive the genetic risk for IPF.

Added value of this study
Using previously identified IPF genetic variants and neuroimaging-derived features from 32,431 participants available through the UK Biobank, we performed the first brain-wide association study for IPF risk variants. We identified brain endophenotypic associations for two IPF risk associated genetic variants, one in chromosome 17 and one in chromosome 8. In particular, increased IPF genetic risk, conferred by a variant near to the *DEPTOR* gene on chromosome 8, was associated with focal cingulate cortical thinning and more widespread changes in white matter microstructure. The cortical association signature was observed in regions with known *DEPTOR* expression while the subcortical findings may be mediated by forced vital capacity (measure of impaired lung function), hinting to distinct direct (cingulate) and indirect (subcortical) brain effects of *DEPTOR* risk alleles.

Implication of all the available evidence
The reported brain-wide IPF risk association patterns reveal two plausible mechanisms that may explain the known association of IPF and brain disorders: (i) direct and focal, thought to be related to paralimbic mTOR dysregulation and (ii) indirect and widespread, in keeping with secondary effects from impaired lung function such as hypoxia. Taken together, these data support the hypothesis that genetic risk profiles may explain some of the observed comorbidity of IPF and brain disorders.

## Introduction

Idiopathic pulmonary fibrosis (IPF) is a serious chronic lung disease of unknown aetiology affecting around five million people worldwide^1^. It is characterised by shortness of breath and causes the lung tissue to become stiff and scar over time. As a result, the lung parenchyma is replaced with a dense extracellular matrix^2^. IPF is typically diagnosed in middle-age, progression and survival is quite variable between affected people^3^, but progressive fibrosis leads ultimately to death with a median survival of 3 to 5 years after diagnosis^4^.

Although the pathogenesis of IPF is incompletely understood, there is a growing body of evidence that multiple risk factors including ageing, genetic alterations, and environmental factors, notably cigarette smoke exposure, contribute to disease risk. Genome-wide association studies (GWAS) have identified a number of loci associated with increased genetic risk in IPF^5^. Although IPF is considered a disease limited to the lung, the mechanisms responsible for the development of fibrosis in the lung are often shared with fibrosis in other organs^6^ which may explain associated comorbidities^7^.

Pulmonary fibrosis has clinically also been associated with cognitive decline^8^, increased risk of neurodegenerative disorders^8^, cerebrovascular accidents^9^, and a high prevalence of anxiety (30-50%) and depression (about 20-30%)^10^. However, the mechanisms linking IPF to brain changes and dysfunction are poorly understood and likely multifactorial; including either direct shared pathomechanisms that result in fibrosis and scar development in the lung and to altered brain development and accelerated neurodegeneration or indirect sequelae of brain injury resulting from fibrosis-associated hypoxic tissue damage. To the best of our knowledge, no post-mortem or in-vivo investigations have been performed to explore these directly.

Neuroimaging can provide unique insight into the nature of the association between IPF and brain dysfunction due to its remarkable sensitivity to subtle structural and functional brain changes. It can reveal topographical and tissue-specific brain signatures linked to genetic risks of primary brain disorders or comorbid brain health in systemic disease. Brain imaging has been successfully used to provide intermediate phenotypes (sometimes referred to as endophenotypes) to assist GWAS studies in brain disorders^11^ or to explore the link between risk and clinical phenotype^12^. Recent advances with population data and tools for systematic extraction of quantitative brain imaging-derived phenotypes (IDPs)^13^ permit the investigation of associations between IDPs and genetic risk profiles to discover novel intermediate brain phenotypes, which may not be directly expressed as behaviours, but highlight altered cellular and molecular functions^11^. This approach is particularly powerful to understand brain comorbidities without robust prior mechanistic or regional knowledge and with low or non-specific clinical manifestations. Therefore, in this study we used a brain-wide exploratory approach, capitalising on population-level datasets available through the UK Biobank^13,14^. Similar to phenome-wide association studies, which examine the associations between specific genetic variants and a wide array of phenotypes across the genome, we performed a brain-wide association study (BWAS) to examine the association of genetic variants linked with the risk of IPF (Suppl. Table S1) across a large array of 1,248 IDPs (Suppl. Table S2), which capture morphological, (micro)structural, and functional brain features.

## Materials and Methods

### Study cohort selection from the UK Biobank

Neuroimaging and genetic datasets from the population-level UK Biobank resource were used^13,14^. From the entire UK Biobank cohort, we derived our study cohort of 32,431 participants (Figure 1), based on *a priori* selection criteria (see Suppl. Material). Briefly, included participants had: i) Neuroimaging (magnetic resonance imaging – MRI) and genetic data; ii) Good-quality anatomical MRI (T_1_-weighted) data; iii) Reliable neuroimage processing and features extracted; iv) White European ethnic background; v) No major neurological conditions reported. To reduce bias from major IDP variation of no interest, we excluded neurological disorders (Multiple Sclerosis, Stroke, Parkinson’s Disease, or Alzheimer’s Disease) with known large effects on imaging features, through the underlying disease or its medication; vi) Available smoking and alcohol status; vii) No relationship to other study participants; viii) Consistent genotype and self-reported sex. Our selected study cohort included individuals aged between 45–82 years (mean=64.2,std=7.5) at the time of imaging.

**Figure 1.**
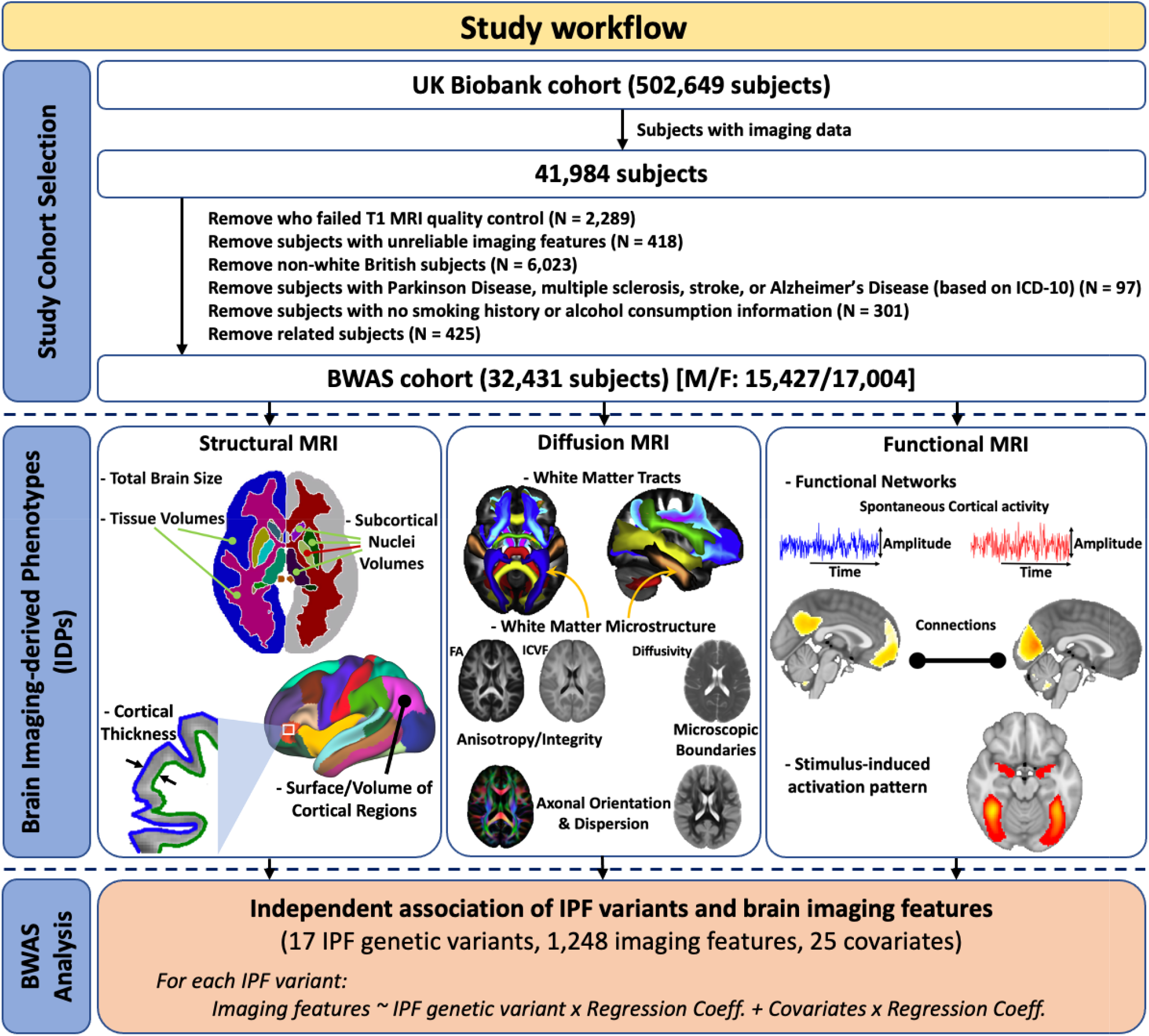
The workflow schematic for data selection, neuroimaging-derived feature extraction, and subsequent BWAS analysis. MRI, magnetic resonance imaging; BWAS, brain-wide association study; IPF, idiopathic pulmonary fibrosis; FA, fractional anisotropy; ICVF, intra-cellular volume fraction; MS, multiple sclerosis.

### Imaging data and imaging-derived phenotypes

A large set of neuroimaging features (also called imaging-derived phenotypes – IDPs) were derived from all available brain imaging modalities provided by the UK Biobank. Modalities included: i) Structural MRI (T_1_-weighted, T_2_-weighted, and susceptibility-weighted imaging (SWI)) that provides anatomical and morphological information; ii) Diffusion MRI (dMRI) that reflects white matter microstructural complexity and integrity derived from two models, the diffusion tensor imaging (DTI) and neurite orientation dispersion and density imaging (NODDI) model; and iii) Functional MRI (fMRI), which indirectly reflects neural activity and cortical connectivity during task-free (resting-state) and task-based conditions. A consistent set of N=1,248 IDPs for each participant, representing a multi-modal brain phenotype (details in Suppl. Material/Table S2), was extracted from the MRI data using a standardised processing pipeline^13,15^ and provided by the UK Biobank.

To exclude effects from imaging confounds (such as image quality, head motion, and head size) that could bias subsequent analysis, we used a set of 45 variables, as identified in^16,17^, to de-confound the IDPs (full list in Suppl. Table S3).

### Genetic data pre-processing

For our study cohort, we used the previously described imputed UK Biobank genotype data released in November 2020 (Data-Field 22828, ver.3 of imputed data)^14^. Seventeen loci have been associated with increased genetic risk in IPF^5,18^ (Suppl. Table S1). All 17 IPF variants (single-nucleotide polymorphisms – SNPs) were present in the UK Biobank imputed data and were used for subsequent analyses.

### Brain-wide association study (BWAS) against IPF variants

We tested the association between each of the 17 IPF variants and each of the 1,248 IDPs across 32,431 individuals. For each IPF variant, separate analyses were performed using a general linear model. An additive genetic effect was assumed (Suppl. Figure S1) and we adjusted for certain covariates, as suggested in^17^: demographic measures (age, sex, age^2^, age × sex, and age^2^ × sex); body measurements (standing height, weight, diastolic and systolic blood pressures, and their squared versions); genetic information (top 10 ancestry principal components, to adjust for population stratification); and lifestyle measures (smoking and alcohol consumption), (see also Suppl. Table S3). The allele associated with increased risk of IPF was chosen as the coded allele. We tested for statistical significance of the observed genetic association after applying Bonferroni correction for multiple comparisons (P-value<0.05/(17×1,248)). The number of subjects with available data for each IDP varied from a minimum of 22,892 to a maximum of 32,431.

### Identification of shared causal variants (co-localisation analysis)

For the genetic variants that were found to be associated with brain IDPs, we performed co-localisation analysis using the COLOC method^19^ (details in Suppl. Material). This allowed us to explore whether the associations between IPF susceptibility (trait 1) with the brain phenotype/IDPs (trait 2) were likely due to the same causal variant (assuming it has been tested and there is only one causal variant). The IPF susceptibility GWAS data were obtained from a large meta-GWAS study of IPF^5^. For the BWAS data, we selected the IDPs that corresponded to the most significant associations (that survived Bonferroni) in BWAS. SNPs within 1□Megabase (Mb) window from each side of the lead SNP were included. For trait 2, we selected the same SNPs (as used for trait 1) within +/-1□Mb window from the lead SNP.

### Mediation analysis

To explore whether associations between neuroimaging features (IDPs) and IPF genetic risk, as derived from the BWAS regression model, were indirect effects from subclinical IPF risk manifestation, we undertook mediation analysis focusing on basic lung function that could plausibly affect brain health. We used forced vital capacity (FVC–best measure, Data-Field 20151), a cumulative measure of lung function (subclinical phenotype)^20^, as an exploratory mediator (Suppl. Material/Figure S2). We constructed a mediation pathway between a single independent variable (IPF genetic risk *X*), a single mediator (FVC *M*), and a single dependent variable (brain IDPs *Y*). This aimed to quantify the direct and indirect relationships between IPF risks, IDPs, and FVC. The direct and indirect standardized effects were calculated using multivariate-adjusted linear regression analysis and a Sobel test^21^ was used to determine whether the indirect effect of the SNP on IDPs through FVC was significant. Only IDPs that were found to associate with potentially causative genetic risk in the previous analyses were used in the mediation models.

### Role of the funding source

Funding sources played no role in study design; collection, analysis, or interpretation of the data; writing of the report, or in submission of this paper for publication.

## Results

### Brain imaging-derived phenotypes associate with IPF genetic variants

We explored brain-wide associations of the 17 IPF genetic variants against 1,248 brain IDPs across 32,431 unrelated white British participants in the UK Biobank study. Figure 2 summarises the results for all 17×1,248 associations. Two of the 17 IPF variants (rs28513081 (G/A) – *DEPTOR* gene in chromosome 8 and rs2077551 (C/T) in chromosome 17) were found to be significantly associated with brain IDPs. The number of associations reaching the Bonferroni-corrected threshold for statistical significance (P-value<2.36×10^−6^) were 19 and 174, respectively for the chromosome 8 and chromosome 17 variants (giving in total 184 brain IDPs – Suppl. Table S4).

**Figure 2.**
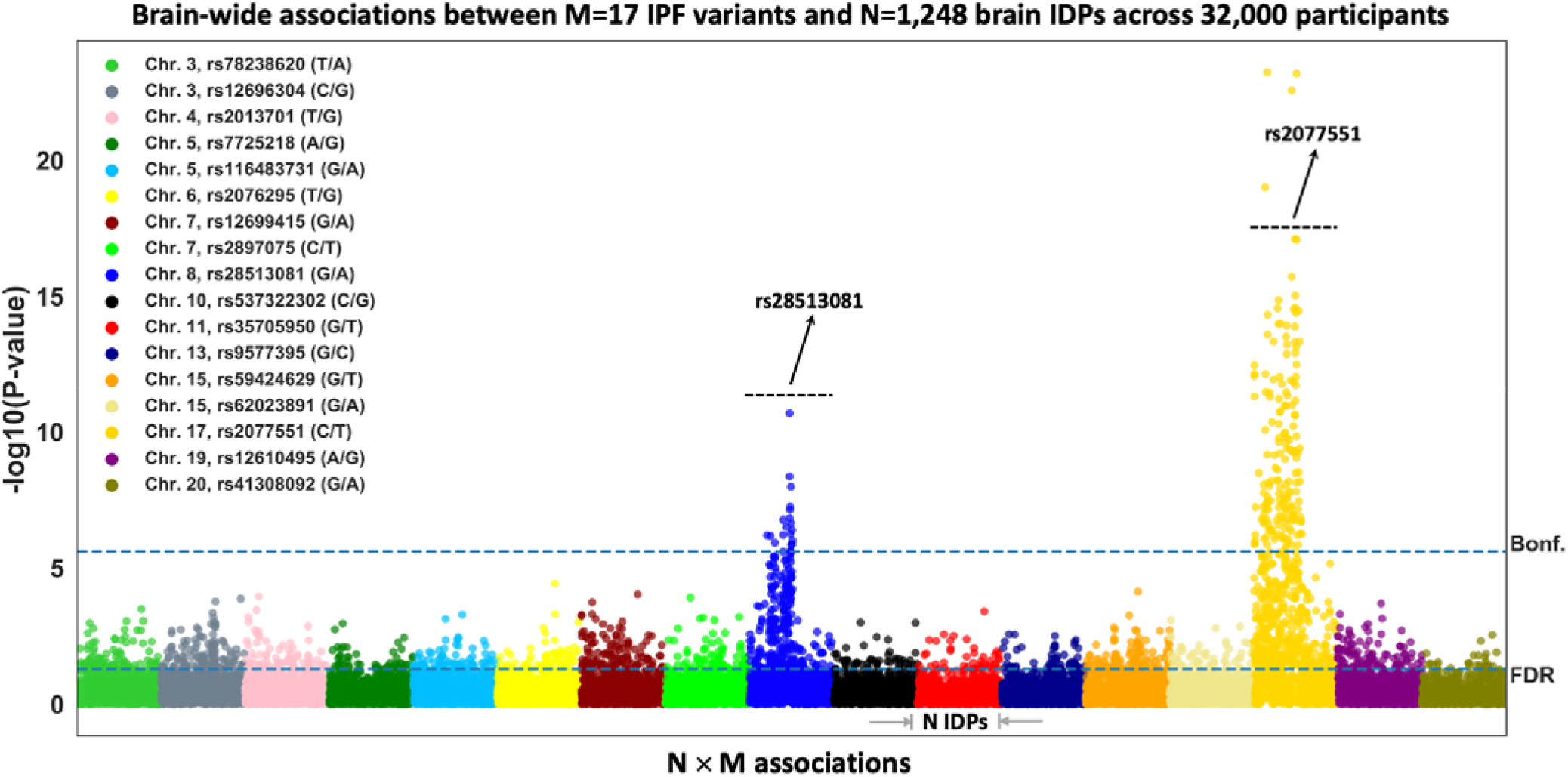
Manhattan plot of the BWAS outcomes of 17 known IPF variants using UK Biobank imaging cohort (S=32,431). Each of the IPF variants is coded by a different colour and within each of these colour groups, associations with the 1,248 brain IDPs, derived from neuroimaging, are depicted. For each association between a SNP with an IDP, its -log10(P-value) is plotted (i.e., M×N=17×1,248 associations are shown). The dashed horizontal line indicates the brain-wide level of significance, i.e., FDR (false discovery rate – bottom line, corresponding to FDR-corrected P-value=0.05) and Bonferroni correction across M×N=21,216 tests (top line, P-value<2.36×10^−6^).

### IPF genetic risk associates with cortical morphology and white matter microstructure

Within the brain IDPs, we explored which categories of features demonstrated associations with the genetic data. For both chromosome 8 and chromosome 17 variants, we found that the associations were driven by brain morphological and microstructural features, in cortex and white matter, and interestingly no associations were found with functional features extracted from functional MRI (e.g., functional networks and connectivity) (Figure 3a and Suppl. Figure S3a).

**Figure 3.**
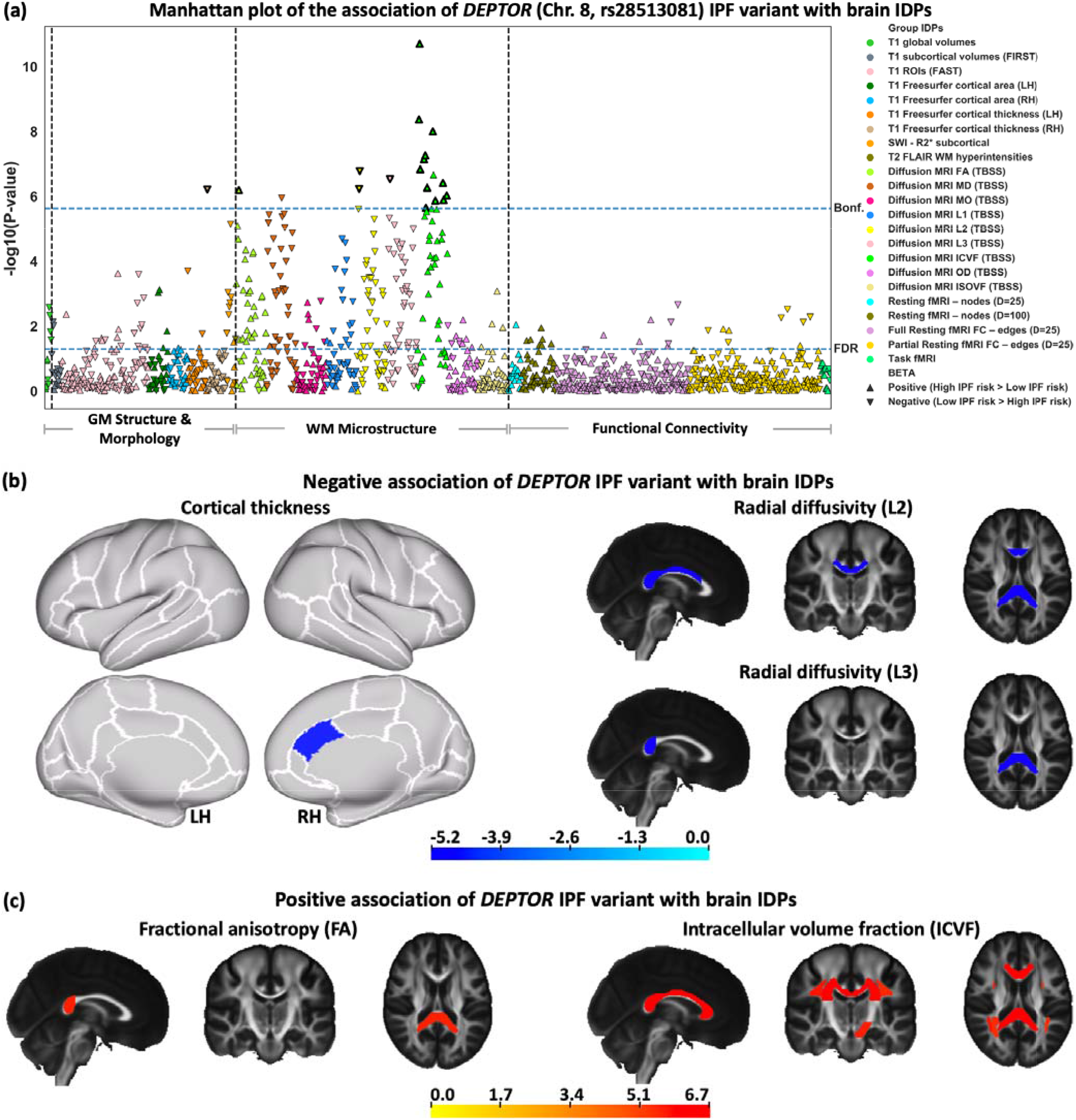
The association of the *DEPTOR* IPF variant with brain IDPs. **(a)** BWAS analysis of rs28513081 (*DEPTOR* gene in chromosome 8). Points are coloured based on the group IDPs. The IDPs included in each group of features are available in Suppl. Table S2. Triangles and inverted triangles are used to show the sign of the BETA in the BWAS. The dashed lines indicate the level of significance, i.e., FDR and Bonferroni thresholds (P-value<2.36×10^−6^). The vertical dashed lines separate the IDPs belongs to the grey matter (GM) structure and morphology, WM microstructure, and functional connectivity. A positive (negative) BETA means that a trait positively (negatively) associated with IPF risk. The triangles with bold outlines in **(a)** demonstrate IDPs that co-localise with IPF susceptibility. (b) Negative and (c) positive associations of neuroimaging features (IDPs) with IPF variant in rs28513081 (*DEPTOR* gene in chromosome 8). T-stat maps of the associations are shown for the most significant IDPs (that survived Bonferroni correction) associated with the fibrosis variant. Positive t-stats correspond to positive association with high IPF risk. T1 Freesurfer-extracted cortical thickness, diffusion MRI L2, and diffusion MRI L3 negatively associated with the *DEPTOR* IPF variant. Diffusion MRI FA and diffusion MRI ICVF positively associated with the *DEPTOR* IPF variant. In the cortical thickness map, the white contours show the outline of the regions in Desikan-Killiany–Tourville (DKT) atlas and top and bottom panels show the lateral and medial view of the brain, respectively. In other spatial maps, the standard HCP1065 FA image is used as background (left is right). LH, Left hemisphere; RH, Right hemisphere.

Figure 3b-c shows spatial maps of the 19 IDPs associated with the *DEPTOR* IPF variant that survived Bonferroni correction, along with the sign and magnitude of the association (t-stat). Cortical thickness in the caudal anterior cingulate was negatively associated with IPF risk (lower thickness↔higher genetic IPF risk). A number of white matter (WM) IDPs (mostly reflecting anisotropy, such as the fractional anisotropy (FA) and the NODDI intracellular volume fraction (ICVF), and radial diffusivity indices) of major fibre bundles (corpus callosum, superior longitudinal fasciculus, and corona radiata) were also associated with the *DEPTOR* IPF variant. These IDPs represent WM integrity and microstructure within these bundles. ICVF and FA correlated positively with IPF risk (Figure 3c), whilst radial diffusivity values correlated negatively (Figure 3b) with high IPF risk (higher ICVF, FA, and lower diffusivity↔higher genetic IPF risk).

We also observed strong associations of brain IDPs with IPF genetic risk for the rs2077551 (chromosome 17) variant, where 174 associations with brain IDPs were found (Suppl. Figure S3). The modalities of associated IDPs and direction of changes were similar to those reported for the *DEPTOR* IPF variant. Significant associations with the rs2077551 (chromosome 17) variant included indices of white matter integrity and microstructure, (such as the fractional anisotropy (FA), the NODDI ICVF, and mean diffusivity (MD)) and morphological measures in the cortex, such as cortical thickness and cortical area, with the strongest association corresponding to “cortical surface area of fusiform – right hemisphere (RH)” (P-value=5.51×10^−24^). No functional measures associated with this IPF variant either. High IPF genetic risk was associated with increased cortical thinning (e.g., inferior and middle temporal regions), with lower diffusivities in white matter (Suppl. Figure S3b), and higher intra-cellular volume fraction/anisotropy in white matter (in bundles including corpus callosum, corona radiata, and association tracts). Conversely to the pattern of associations identified with the *DEPTOR* variant, positive associations of cortical surface area with the chromosome 17 IPF variant were also found (e.g., fusiform and lingual areas) (Suppl. Figure S3c).

To explore whether the association of IPF genetic risk with the brain IDPs was likely due to the same causal variant, we performed co-localisation analysis for the IDPs that were found to be significantly associated with IPF variants. Seventeen of the nineteen significant IDP associations for the chromosome 8 IPF locus showed statistical evidence through co-localisation of a likely shared causal variant with the IPF association (Figure 4, posterior≥0.8^22^ and Suppl. Table S5). For the chromosome 17 locus, co-localisation analysis was inconclusive due to the complex structural variation in the region that results in a large number of highly correlated SNPs across 1.5 Mb^23^.

**Figure 4.**
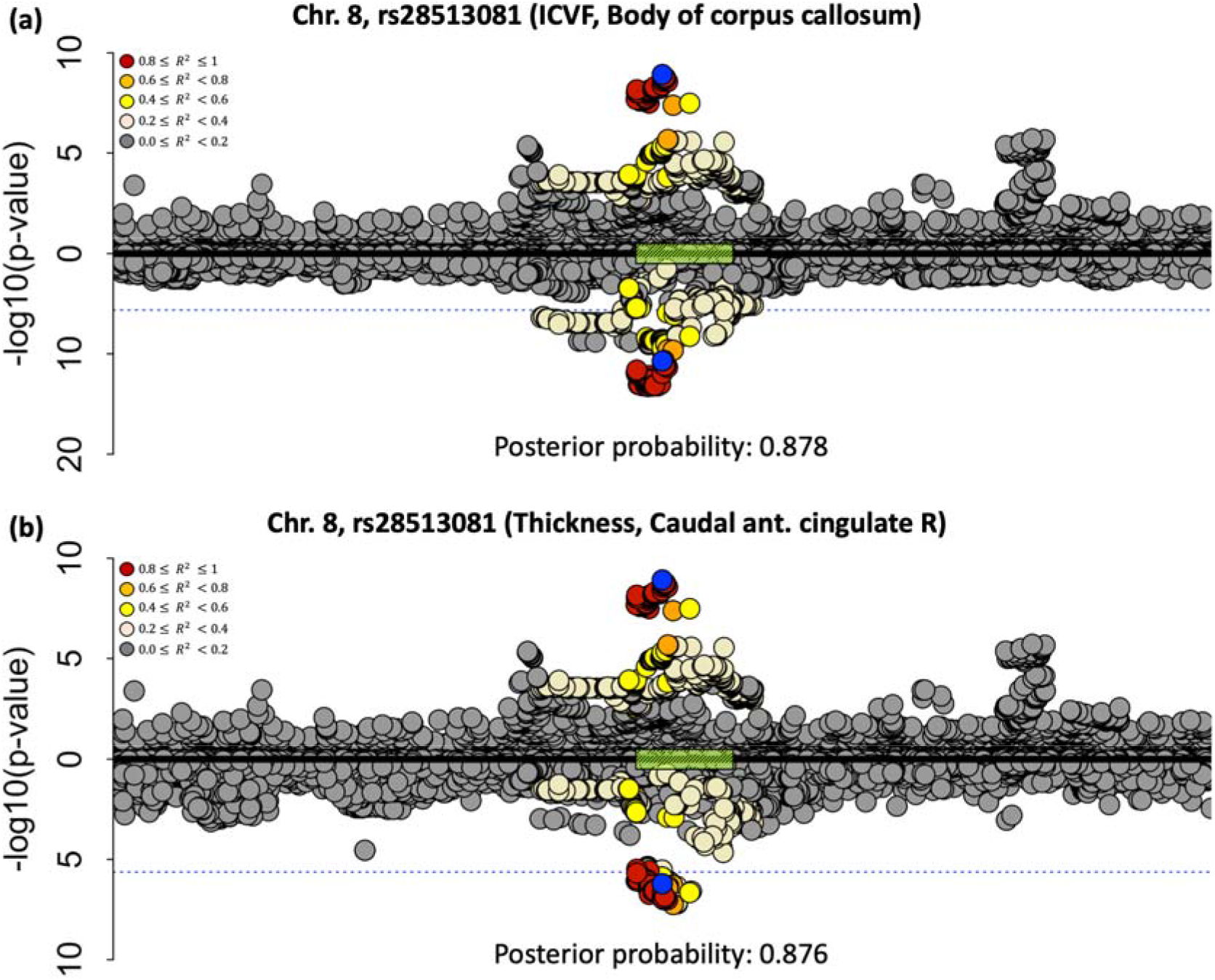
Co-localisation of the two most significantly associated IDPs with chromosome 8 (region around the *DEPTOR* gene), against IPF risk. **(a)** Against a white matter microstructure feature (ICVF of the body of corpus callosum), **(b)** Against a cortical feature (cortical thickness of the caudal anterior cingulate RH). Chromosome 8: 119,934,133–121,934,069 build x-axis for both plots. Each point represents a genetic variant with chromosomal position on the x-axis and –log(P-value) on the y-axis. The GWAS data for IPF risk is presented above the x-axis, and the GWAS data for the IDP is shown below the x-axis. The sentinel variant from the IPF GWAS is shown in blue and other variants are coloured by their linkage disequilibrium with the IPF GWAS sentinel. The dashed blue lines indicate the Bonferroni threshold (2.36×10^−6^). The green box on the x-axis demonstrates the position of the *DEPTOR* gene.

Taken together, the above results provide for the first time strong evidence – through complementary analyses – of a potentially causal association between the *DEPTOR* (chromosome 8) IPF variant and 17 brain IDPs, representing cortical morphology and white matter microstructure. We kept these 17 IDPs for subsequent analysis.

### Associations of brain IDPs with the DEPTOR IPF variant show a dual effect, partially mediated by lung function

To explore potential mechanisms underlying the identified associations, we conducted a mediation analysis. Given that IPF is a restrictive lung disease characterised by impaired lung capacity, we used forced vital capacity (FVC), which is used for IPF diagnosis and characterisation^24^, to examine whether the associations between genetics and neuroimaging features are mediated through variation in lung function irrespective of underlying pathology. We hypothesised that the effect of IPF genetic risk on brain IDPs is indirect via its influence on FVC (Suppl. Figure S2).

Mediation analysis suggested a partial mediation of the *DEPTOR*–IDP association through lung function (FVC, Figure 5a, full results in Suppl. Figure S4/Table S6) with a number of IDPs (5 IDPs, all representing white matter microstructure in the corpus callosum, cerebral peduncles, and thalamic radiations) revealing a significant effect through the indirect FVC mediated pathway. For these associations, the direct path was attenuated but remained significant in line with partial mediation by FVC. As an example, the details of direct and indirect paths of a positively and a negatively associated IDP are shown in Figure 5a. Interestingly only subcortical (white matter) brain IDPs showed FVC mediation effects of their associations with IPF genetic risk suggesting a tissue-type specific partial mediation.

**Figure 5.**
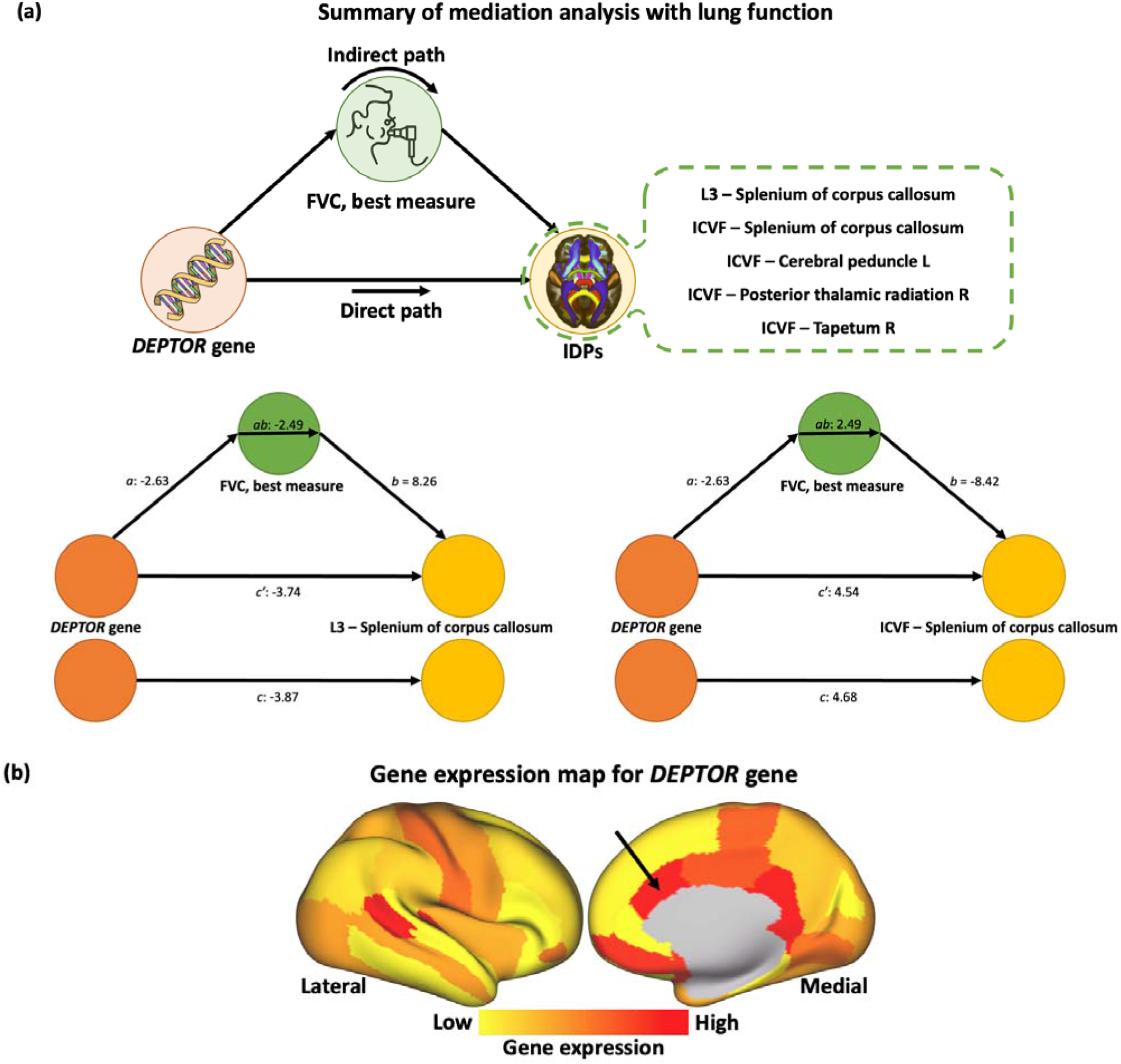
Secondary analysis. **(a)** List of the brain IDPs that associate with *DEPTOR* gene via FVC as a mediator and two examples of the mediation analyses results. The plots demonstrate the direct, indirect (mediated by FVC – best measure), and total effect paths from the *DEPTOR* gene to brain IDPs. The bottom left path diagram in **(a)** shows the mediation through FVC for *DEPTOR* which is negatively associated with the diffusion MRI L3 (splenium of corpus callosum). The bottom right path diagram in **(a)** depicts the mediation through FVC for *DEPTOR* which is positively associated with the diffusion MRI ICVF (splenium of corpus callosum). The Z-scores of the regression coefficient are shown for paths *a, b, c, ab*, and *c’*. **(b)** The group-level gene expression patterns for chromosome 8 (*DEPTOR* gene) projected on an inflated cortical surface. The colour bar shows the gene-expression level. Red regions show areas where the gene of interest is highly expressed, whereas yellow regions indicate low expression values. The black arrow denotes the caudal anterior cingulate, where reduced cortical thickness has been found to associate with the IPF *DEPTOR* variant.

Our mediation results suggest a second distinctly different mechanism that lacks FVC mediation, hinting to a likely direct gene effect that is limited to the anterior caudal cingulate cortex. To corroborate the likely direct nature of this effect, we computed a bulk gene expression map of the *DEPTOR* gene, using the Allen Human Brain Atlas (AHBA) database^25^ and the same cortical surface parcellation as the one used in our IDP extraction pipeline (details in Suppl. Material). As shown in Figure 5b, the caudal anterior cingulate region (denoted by an arrow), where reduced cortical thickness was found to associate with high IPF genetic risk (as captured by the *DEPTOR* IPF variant), has a relatively high expression level for the *DEPTOR* gene. That provides face validity evidence that the observed association between IPF risk gene variant and IDP changes was detected in a cortical region where the risk gene is indeed strongly expressed.

## Discussion

We undertook a large brain-wide multi-modal imaging and genetic association study in 32,000 participants of the UK Biobank to establish brain endophenotypes of IPF genetic susceptibility for the first time. Strong associations with a likely shared causal variant were found for several of the 1,248 studied brain imaging-derived phenotypes (IDPs) for two of the 17 IPF risk associated variants on chromosomes 8 and 17. Co-localisation provided additional evidence that the associated neuroimaging features and IPF share a single causal variant at the chromosome 8 locus. The association pattern consists of focal cingulate cortical thinning and more widespread microstructural impairment in major white matter tracts, such as the corpus callosum and corona radiata. Furthermore, we suggest potential partial mediation of the subcortical IDP–*DEPTOR* variant associations by lung function in keeping with an indirect effect such as systemic hypoxia or inflammation. Conversely, the cingulate thinning was independent of lung function suggesting a direct effect from *DEPTOR* variant expression in view of the known strong *DEPTOR* gene expression in the anterior cingulate^26^.

*DEPTOR* on chromosome 8 encodes for DEPTOR (DEP [dishevelled, Egl-10 and Pleckstrin] domain-containing mechanistic target of rapamycin [mTOR]-interacting protein), which is a key modulator (partial inhibitor) of the mTOR pathways interacting with both of its complexes (mTORC1 and mTORC2)^27^, such that higher levels of *DEPTOR* lead to decreased mTOR activity^28^. In just over a decade since the discovery of *DEPTOR*, major roles have already been established in cancer, metabolism, and immunity explained by its ubiquitous tissue expression and modulation of fundamental cellular processes. In IPF, the recently identified *DEPTOR* IPF risk allele together with decreased gene expression in lung tissue^5^ highlights a risk mechanism through the induction of profibrogenic phenotypes linked to mTORC1 signalling^29^. Interestingly, *DEPTOR* inhibition was also found in brain tissue in Alzheimer’s Disease^30^ and the *DEPTOR*/mTOR ratio is considered to regulate the neuroprotection/neurodegeneration balance in pro-inflammatory states via autophagy regulation. Moreover, *DEPTOR* central nervous system (CNS) expression in the hypothalamus, circumventricular organs, and autonomic nervous regions support increasing evidence for a further critical role in brain-body homeostatic control^26^.

Our analysis provides exciting novel insights that *DEPTOR* dysregulation through the IPF risk variant is linked with cortical thinning and microstructural changes of major white matter tracts. Moreover, we were able to identify possible direct *DEPTOR* and indirect lung phenotypic effects using forced vital capacity (FVC), as established lung function marker in IPF^31^, and bulk gene expression data. The observed IPF risk variant association with distinct cortical thinning in the caudal anterior cingulate did not show FVC mediation and is spatially co-localised with high *DEPTOR* bulk expression in the Allen Human Brain Atlas^25^. This may provide a molecular mechanism to explain the clinical observation of CNS comorbidities in IPF, especially the common anxiety, depression^10^, and cognitive decline^8^. It is conceivable that the IPF risk *DEPTOR* variant will also reduce *DEPTOR* expression in areas of high expression such as the anterior cingulate resulting in the observed cortical thinning. The affection of the anterior cingulate in turn may well explain the development of neuropsychiatric comorbidities as evidenced by a large body of neuroimaging studies^32^.

We propose a second indirect mechanism for the IPF genetic risk associations with more widespread changes in major white matter tracts with potential mediation through FVC. The specific characteristics of these imaging associations point to a neuroinflammatory signature that may relate to chronic systemic pro-inflammatory state or tissue hypoxia. High ICVF and reduced diffusivity measures that were correlated with high IPF risk correspond to findings in acute neuroinflammation and injuries^33^, such as traumatic brain injury (TBI), axonal injury, and possible hypoxic swelling. Even if some mechanisms may be shared (e.g., microglial activation), it is unlikely that the white matter microstructure patterns we observe here reflect an acute mechanism of inflammation, but instead, we propose chronic low-level neuroinflammation combined with reduced tissue oxygen levels. Animal models of stress/hypoxia have shown reduced diffusivity in WM (e.g., increased fractional anisotropy and reduced mean and radial diffusivity after 2 weeks of stress^34^ reduced diffusivity values in early hypoxia^35^). Notably, reductions of (radial) diffusivity can lead to apparent increases in ICVF, as estimated by the NODDI model^36^. This model assumes constant diffusivities, so ICVF can be overestimated when diffusivity (assumed constant in the NODDI model) is lower than assumed. Taken together, hypoxia and chronic neuroinflammation are plausible mechanisms for the observed pattern of subcortical changes in diffusivity.

We detected conspicuously strong effects of IPF genetic risk on brain structure for chromosome 17 that are noteworthy despite the inconclusive co-localisation analysis. The subcortical endophenotypic pattern of the chromosome 17 IPF risk variant was broadly similar to the microstructural changes seen for the *DEPTOR* risk variant and also overlaps spatially suggesting a similar indirect pathomechanism warranting further mechanistic and mediation studies. Interestingly, the cortical chromosome 17 risk variant signature showed a distinct spatial profile and displayed a dichotomy with cortical thinning but increased cortical surface area. Such a pattern has been reported in some neurodegenerative conditions like Parkinson’s Disease^37^ and points to shape variation such as increased gyrification. The nature of these dissociated cortical changes remains unknown but has been linked to putative neurodevelopmental differences^38^ as cortical thickness and area show very little genetic correlation^39^.

A limitation in our study is the lack of prior evidence that *DEPTOR* gene expression levels change in the anterior cingulate cortex in association with the IPF risk variant, revealed by absence of a relevant expression quantitative trait loci (eQTL) signal in the Genotype-Tissue Expression (GTEx) portal. Whilst this may counter the hypothesis that there is a direct effect of *DEPTOR* on cortical thickness, the GTEx brain cortex analysis may not have sufficient power to identify an effect, as brain cortex samples were amongst the smallest compared with other tissues profiled in GTEx (https://gtexportal.org/home/tissueSummaryPage#donorInfo). A further limitation is that our BWAS analysis has not accounted for the likely covariances between non-orthogonal IDPs. We have however used a stringent Bonferroni threshold for correction of multiple comparisons for primary outcomes, hence the identified associations are likely conservative. No multiple test correction was used for the mediation analysis for which we used an absolute FVC measure (in litres), however we have accounted in the mediation for age, sex, height, and ethnicity (our study cohort is restricted to white European ethnic background).

In summary, we exploited imaging and genetic data from more than 32,000 participants available through the UK Biobank population-level resource to explore links between IPF genetic risk and imaging-derived brain endophenotypes for the first time. We identified strong associations between cortical thickness and white matter microstructure and IPF risk loci in chromosome 8 (*DEPTOR gene*) and chromosome 17. Through co-localisation a shared causal gene locus was identified for the *DEPTOR* variant and associated brain signature, and for cingulate cortical thinning no mediation effect from lung function was found. Taken together, these data support the hypothesis that genetic risk profiles may explain some of the observed comorbidity of IPF and brain disorders with further mechanistic studies warranted to characterise additional indirect effects.

## Data Availability

All data used in this study are publicly available from the UK Biobank (www.ukbiobank.ac.uk). Our study was performed under UK Biobank Project 43822.

## Acknowledgements

Data were provided by the UK Biobank under Project ID 43822 (PI: Sotiropoulos). This study was supported by the NIHR-funded Nottingham Biomedical Research Centre and the DEMISTIFI consortium – (Medical Research Council grants MR/V005324/1, MR/W014491/1). RA is an Action for Pulmonary Fibrosis Research Fellow. LMK was funded by a Medical Research Council PhD studentship (MR/N013913/1). LVW holds a GSK / Asthma+Lung UK Chair in Respiratory Research (C17-1). SS is supported by a Consolidator Grant from the European Research Council (ERC/101000969). The computations described in this paper were performed in part using the University of Nottingham’s Augusta HPC service and the Precision Imaging Beacon Cluster, which provide High Performance Computing service to the University’s research community. The research was partially supported by the NIHR Leicester Biomedical Research Centre; the views expressed are those of the author(s) and not necessarily those of the National Health Service (NHS), the NIHR, or the Department of Health.

## Declaration of interests

RGJ reports research funding from AstraZeneca, Biogen, Galecto, GlaxoSmithKline, RedX, Pliant, and Genetech; personal fees from Bristol Myers Squibb, Daewoong, Veracyte, Resolution Therapeutics, RedX, Pliant, Chiesi, Roche, PatientMPower, AstraZeneca, GSK, Boehringer Ingelheim, Galapagos, and Vicore; non-financial support from NuMedii and Action for Pulmonary Fibrosis, outside the submitted work. LVW reports research funding from GSK and Orion Pharma and have conducted consultancy for Galapagos. DPA reports research funding from Biogen.

All other authors declare no conflict of interest.

## Author’s contributions

AM: data curation, methodology, formal analysis, writing – original draft, RJA: methodology, formal analysis, writing – review & editing, LMK: methodology, writing – review & editing, OCL: resources, writing – review & editing, RGJ: conceptualisation, funding acquisition, writing – review & editing, LVW: conceptualisation, methodology, funding acquisition, writing – review & editing, DPA: supervision, conceptualisation, funding acquisition, writing – original draft, SNS: methodology, resources, supervision, funding acquisition, conceptualisation, writing – original draft.

## Notes

### Author Declarations

The study used (or will use) ONLY openly available human data that were originally located at the UK Biobank data

