## Supplementary Materials for "Mapping brain endophenotypes associated with idiopathic pulmonary fibrosis genetic risk"

### Supplementary Text

Filtering criteria used to extract study cohort 3

Imaging data and imaging-derived phenotypes (IDPs) 3

IDP De-confounding 4

BWAS Covariates 5

The COLOC algorithm 6

Mediation analysis 6

Allen Human Brain Atlas gene expression cortical maps 6

References 8

### Supplementary Tables

Table S1. List of the genetic variants representing the 17 reported IPF susceptibility signals. 10

Table S2. List of all imaging-derived phenotypes (IDPs) used in this study. 11

Table S3. List of confounds/covariates used in BWAS analysis. 12

Table S4. List of all significant IDPs associated with IPF risk genetic variants. 174 IDPs associated with the Chr. 17 (17q21.31 gene) IPF variant and 19 IDPs associated with the Chr. 8 (DEPTOR gene) IPF variant. 13

Table S5. Co-localisation (COLOC) posterior probabilities (H0 – H4) for IDPs that were associated with Chr. 8 (DEPTOR gene) variant and Chr. 17 (17q21.31 gene) variant in the BWAS model. 16

Table S6. Mediation analysis coefficients, of DEPTOR variant to IDP association, through lung FVC. 17

### Supplementary Figures

Figure S1. The general linear model that is used in BWAS analysis 18

Figure S2. Schematic of a simple mediation model. 19

Figure S3. The association of chromosome 17 IPF variant with brain IDPs. 20

Figure S4. Summary of mediation analysis for the 3 (out of 5) IDPs with significant mediation through FVC, not shown in Main text. 22

### Supplementary Text

#### Filtering criteria used to extract study cohort

From the full UK Biobank cohort (~500,000 participants), we considered participants that i) Had neuroimaging data available. Based on the UK Biobank data released in April 2020, only 41,984 subjects had brain imaging data. ii) Had good quality T1w magnetic resonance imaging (MRI) data. We excluded 2,289 subjects because of poor structural imaging data (for example, due to having incomplete brain coverage, or having very severe MRI artefacts)[1]. iii) Had good quality neuroimage processing and features extracted. Imaging-derived feature (IDP) values that were greater/less than five times the standard deviation from the cohort-mean IDPs were considered technical outliers and thus removed. We removed 418 subjects due to unreliable imaging features in more than 10 IDPs. iv) Had white European ethnic background, to keep ethnicity consistent. Based on the self-reported ethnicity or the similarity of their genetic ancestry, 6,023 participants were non-white European and were excluded (Data-Field 22006). v) Did not have potentially confounding neurological conditions. Based on the main and secondary ICD-10 (Data-Field 41202 and 41204), 97 individuals were excluded from the analysis due to been diagnosed with either Parkinson's Disease (code G20, G21, G22, G210 – G214, G218, G219), Multiple Sclerosis (code G35), Alzheimer's Disease (code G30, G300, G301, G308, G309), or had a Stroke (code G46, G460 – G468, I60 – I64, I600 – I616, I618 – I621, I629 – I636, I638 – I639). vi) Had available information on smoking status and alcohol drinking. We excluded 301 individuals that did not have smoking status (Data-Field 20116), alcohol drinker status (Data-Field 20117), or alcohol intake frequency (Data-Field 1558) information available. vii) Were unrelated to other study participants. We removed 425 subjects that were family-related to other individuals in the cohort with a KING kinship coefficient ≥ 0.0884[2] (from each group of related individuals we chose only one representative, forming a maximally unrelated subset – relatedness file downloaded from UK Biobank). viii) Their genotype and self-reported sex matched (Data-Field 31 and 22001).

#### Imaging data and imaging-derived phenotypes (IDPs)

The neuroimaging data consisted of five MRI modalities (T1-weighted, T2-weighted, susceptibility-weighted, diffusion MRI, and functional MR). From these, three main categories of features (IDPs) could be extracted, structural, microstructural, and functional, totalling 1,248 IDPs.

The structural IDPs provide information about morphological and anatomical aspects of the brain's macroscopic structures. Features include: i) T1-weighted: Sensitive markers of atrophy, which can be both global and local, e.g., total brain size, tissue volumes (grey matter (GM), white matter (WM), and cerebrospinal fluid (CSF)), volumes of subcortical GM (e.g., thalamus and hippocampus), volumes of cortical/cerebellar regions, and Freesurfer-extracted[3,4] cortical surface area and cortical thickness. ii) T2-weighted: provide information on T2-hyperintensities, which represent WM lesions. iii) susceptibility-weighted imaging (SWI):

Features are iron content proxies in subcortical structures (R2* values) and can reflect iron deposition associated with neurodegeneration.

Microstructural IDPs are derived from diffusion MRI and are proxies of tissue complexity and integrity in white matter. They have been estimated using two microstructural models, the diffusion tensor imaging (DTI) and neurite orientation dispersion and density imaging (NODDI) models and have been averaged in WM regions of interest (ROIs), representing 48 white matter tracts. The features we considered were DTI fractional anisotropy (FA), DTI mean diffusivity (MD), DTI mode of anisotropy (MO), and DTI eigenvalue maps (L1, L2, and L3), NODDI intra-cellular volume fraction (ICVF, an estimate of neurite – i.e., axons and dendrites – density), NODDI orientation dispersion (OD, which expresses the extent of complexity of fibre orientations), and NODDI isotropic volume fraction (ISOVF, an index of the relative extra-axonal water diffusion).

Finally, functional IDPs features are derived from functional MRI and indirectly reflect neural activity. Functional MRI (fMRI) measures the dynamic changes in blood oxygenation and flows which is the result of changes in neural metabolic demand. UK Biobank has both resting-state (task-free) fMRI (rfMRI) and task fMRI (tfMRI) data. In our analysis, we used the regional resting-state activity peak amplitude (a marker of overall spontaneous activity for prespecified spatially independent brain regions – networks) and resting-state network functional connectivity (FC) indices, that reflect synchrony in spontaneous regional activity. The connectivity estimates are available for both coarse (D = 25) and fine (D = 100) region definitions. From the task fMRI data, we considered features that represent activity in response to an external stimulus (emotional task). A list of all features considered as IDPs is shown in **Table S2**.

#### IDP de-confounding

As imaging-derived features can be affected by non-biological parameters, we de-confounded them following the recommendations in[5,6]. In total, 45 features were used as the main confounds set for this study. The confound variables included: a) Volume-to-volume head motion, the micro-movements of the head between two consecutive volumes (time-points) in dynamic imaging (rfMRI and tfMRI), averaged across all volumes. b) Head size scaling, defined as the volumetric scaling from the T1 head image to a template MNI152 standard space image. c) Head position within the scanner, described by the scanner lateral (X), transverse (Y), and longitudinal (Z) brain position, and the scanner table position, as data quality is highly dependent on the exact location of the head and the radiofrequency receive coil in the scanner. d) The MRI scanning site, as the data have been collected from three UK Biobank imaging sites. Even if UK Biobank uses identical scanner hardware and software for all three sites, there are still subtle differences between them. e) Date-related drift. As suggested by[5,6], there are slowly changing drifts in the data. To capture those, we first generated a matrix of Subjects × IDPs. To overcome any missing data, this matrix was imputed using a low-rank matrix imputation algorithm[7]. To remove outliers, we applied a median-based outlier removal (discarding values greater than five times the median absolute deviation from the overall median). The matrix was then temporally regularized (with respect to the scan date – Data-Field 53) with spline-based smoothing. Then, after a PCA, the top 10 components were kept to reflect the primary modes of slowly changing drifts in the data and be used as confounds.

As suggested in[5,8,9], for these confounding variables we generated augmented versions as well (for example quantile normalisation, outlier removal, and centred squared versions of these confounds). **Table S3.** provides a list of all the confounding variables which was used. A general linear model of the original IDPs against these confounding regressors allowed us to regress out their effects on the IDPs and obtain de-confounded IDPs.

#### Brain-wide association study (BWAS) covariates

The de-confounded IDPs were used along with single-nucleotide polymorphism (SNP) data in the BWAS model to identify brain-wide Imaging-Genetics associations. We used the following variables as covariates in the BWAS model: i) Demographics. we used age (the difference between the date of birth and scanned date, Data-Fields 34, 52, and 53), sex (Data-Field 31), $age^2$, age × sex, $age^2$ × sex.  ii) Body measurements. We used standing height (Data-Field 50), and weight (Data-Field 21002) as covariates. Any missing value in height and weight data was substituted by the imputation (using the R package "mice"[10]) of height and weight values prior to imaging stages and the second imaging session (if available). Diastolic and systolic blood pressures were measured several times during the imaging visit using automated and manual blood pressure reading devices (Data-Fields 4079, 4080, 79, 80). We averaged the automated and manual blood pressure readings. The missing values were replaced with the imputed version of these readings from other sessions. iii) Genetic variables. We used genetic matrix principal components (the top 10 principal components, Data-Field 22009), as covariates, iv) Lifestyle measures. Measures related to smoking and alcohol consumption were used as covariates in the model. For smoking, we generated an 'ever smoker' covariate. To derive this covariate, we used 'smoking status' (Data-Field 20116). This field summarises the current/past smoking status of the participants. For the participants who have clearly answered this question at the imaging visit, that answer is used for their status. Otherwise, we used their answer for their first repeat assessment or the initial assessment visit. Then, previous and current smokers were grouped together to make an 'ever smoker' covariate (1: current/previous, 0: never). For alcohol consumption, we derived a variable that reflects the number of alcohol units per week for each participant. To do that, for all participants who answered they consumed alcohol (Data-Field 20117), we used their alcohol intake frequency (Data-Field 1558) and their intake for different types of drink per week or per month using standard drink sizes (Data-Fields 1568, 1578, 1588, 1598, 1608, 5364, 4407, 4418, 4429, 4440, 4451, 4462). Then, for each type of drink, we applied a standardized number of UK alcohol units to be able to estimate the number of units of alcohol consumption per week for each participant[11].

The covariates were de-meaned and unit-variance normalised. In total, 25 variables were used as covariates in the BWAS models (**Table S3**). When multiple instances of a covariate were available, the values recorded at the imaging visit were used.

#### The COLOC algorithm

The COLOC algorithm was used to explore whether the associations between idiopathic pulmonary fibrosis (IPF) susceptibility (trait 1) and with the brain phenotype/IDPs (trait 2) were likely due to the same causal variant. This algorithm utilizes an approximate Bayesian factor to estimate the posterior probability of each of the following models: (i) There is no association in the genome region for traits 1 or 2 (PP.H0). (ii) There is an association in the region with trait 1 but not trait 2 (PP.H1). (iii) There is an association in the region with trait 2, but not trait 1 (PP.H2). (iv) There is an association in the region with both traits 1 and 2, but these are driven by two distinct causal sources (PP.H3). (v) There is an association in the region with both traits 1 and 2, driven by the same causal source (PP.H4). PP.H4 indicates shared overlap between the traits. Co-localisation results were plotted using R (version 3.5.1) and the Mirrorplot package (https://github.com/rjallen513/mirrorplot).

#### Mediation analysis

As described in Figure S2, we ran a mediation analysis to check whether the association between the *DEPTOR* gene IPF variant $X$ and the brain IDPs $Y$ was mediated through forced vital capacity (FVC) $M$, a measure of lung function. A significant mediation was identified when: 1) The indirect ($X \rightarrow M \rightarrow Y$) and direct ($X \rightarrow Y$) pathways were governed by significant associations, i.e., coefficients $a$, $b$, and $c'$ were statistically different than zero. A Sobel test was used to check whether $ab$ was different than zero. 2) The association between $X \rightarrow Y$ was attenuated when the mediator $M$ was included in the regression model, i.e., $|c'| < |c|$, indicative of partial mediation. We performed mediation analysis using the M3 package[12]. All associations were calculated by controlling for the same potential covariates as described above. Supplementary Table S6 contains the results

#### Allen Human Brain Atlas gene expression cortical maps

We generated cortical maps of bulk gene expression for specific genes relevant to IPF variants, using data from the publicly available Allen Human Brain Atlas (AHBA)[13,14]. Gene expression data, as measured from spatially distinct histologically validated neuroanatomical tissue samples (from cortical, subcortical, brainstem, and cerebellar regions) using microarrays, consisted of multiple genetic probes. We considered tissue samples from the left hemisphere (as suggested from their standard space coordinates) of six neurotypical adult brains (five male and one female aged 24–57 with a mean age of 42 years)[13]. These data were further processed to generate whole-brain gene expression maps, for specific genes.

For a gene of interest GID, such as the *DEPTOR* gene, the followed procedure was as follows: 1) Probe-to-gene re-annotation and selection of gene probes with a valid GID. The AHBA provides annotation tables that shows which gene each probe refers to. Updates of the sequencing databases gets this information outdated[15]. We used a re-annotator toolkit[16] to re-annotate probes and select the ones relevant to GID. 2) Data filtering. The AHBA provided a binary indicator for each probe showing that the measured expression level exceeds well the background level. We used this binary indicator as quality control to exclude probes for which the measured expression levels were below the background[14]. 3) Probe selection. Multiple probes can refer to the same gene. Due to errors that arise from different sources [for further details see[17]] inconsistent expressions can arise between these probes. We selected the probe with the most consistent pattern of regional variations across the six donor brains for the specific gene GID[14,15]. 4) Assign probe locations to brain regions. First, for each gene, the gene expressions were Z-scored. For each subject, for each tissue sample, we created a sphere whose centre is the MNI coordinate of that tissue sample with a radius of 3 mm. Finally, the gene expression levels detected by each probe in each sample was assigned to all of MNI voxels within this sphere. Adding all tissue samples and gene expression measurements for a gene gave a volumetric gene expression map for each subject. 5) Project volumetric gene expression maps to the cortical surface. For each subject, the gene expression maps were projected from volumetric space onto cortical surface meshes. The Z-scored gene expression surface maps of all the subjects were then added together. Based on the Desikan-Killiany-Tourville (DKT)[18] parcellation atlas (34 parcels per hemisphere), a parcellated group-level gene expression surface map was obtained for GID, with the value in each parcel being the median of non-zero gene expression values within that parcel.

For the *DEPTOR* gene, we selected probes A_23_P60166 and A_24_P224488 from the AHBA database, as they had the most consistent patterns of regional variations across the six donor brains.

### Supplementary Tables

**Table S1.** List of the genetic variants representing the 17 reported IPF susceptibility signals.

| Chr. | Position | rsid | Locus | Ref allele | IPF risk increasing allele | Risk allele frequency |
|---|---|---|---|---|---|---|
| 3 | 44902386 | rs78238620 | *KIF15* | T | A | 5.3% |
| 3 | 169481271 | rs12696304 | *LRRC34/TERC* | C | G | 27.9% |
| 4 | 89885086 | rs2013701 | *FAM13A* | T | G | 51.3% |
| 5 | 1282414 | rs7725218 | *TERT* | A | G | 67.5% |
| 5 | 169015479 | rs116483731 | *SPDL1* | G | A | 0.8% |
| 6 | 7563232 | rs2076295 | *DSP* | T | G | 46.9% |
| 7 | 1909479 | rs12699415 | *MAD1L1* | G | A | 42.0% |
| 7 | 99630342 | rs2897075 | *7q22.1* | C | T | 39.1% |
| 8 | 120934126 | rs28513081 | *DEPTOR* | G | A | 57.2% |
| 10 | 93271016 | rs537322302 | *HECTD2* | C | G | 0.3% |
| 11 | 1241221 | rs35705950 | *MUC5B* | G | T | 14.9% |
| 13 | 113534984 | rs9577395 | *ATP11A* | G | C | 79.3% |
| 15 | 40720542 | rs59424629 | *IVD* | G | T | 53.9% |
| 15 | 86097216 | rs62023891 | *AKAP13* | G | A | 30.0% |
| 17 | 44214888 | rs2077551 | *17q21.31* | C | T | 81.4% |
| 19 | 4717672 | rs12610495 | *DPP9* | A | G | 30.5% |
| 20 | 62324391 | rs41308092 | *RTEL1* | G | A | 2.1% |

**Table S2.** List of all imaging-derived phenotypes (IDPs) used in this study. The 1,248 IDPs are grouped into categories based on MRI modality and type of information they encode.

| Group IDP Name | Description | Number of IDPs | UK Biobank Data Field |
|---|---|---|---|
| **T1 global volumes** | Contains features like total brain size, and normalised/unnormalized tissue volumes (GM, WM, and CSF) (based on SIENAX) | 11 | 25000 – 25010 |
| **T1 subcortical volumes (FIRST)** | Contains the volumes of subcortical GM (based on FIRST) | 15 | 25011 – 25025 |
| **T1 ROIs (FAST)** | Contains the volumes of cortical/cerebellar regions (based on FAST) | 139 | 25782 – 25920 |
| **T1 Freesurfer cortical area (LH)** | Cortical surface area measure for 31 ROIs (LH) based on the Freesurfer analysis | 31 | 27143 – 27173 |
| **T1 Freesurfer cortical thickness (LH)** | Cortical thickness measure for 31 ROIs (LH) based on the Freesurfer analysis | 31 | 27174 – 27204 |
| **T1 Freesurfer cortical area (RH)** | Cortical surface area measure for 31 ROIs (RH) based on the Freesurfer analysis | 31 | 27236 – 27266 |
| **T1 Freesurfer cortical thickness (RH)** | Cortical thickness measure for 31 ROIs (RH) based on the Freesurfer analysis | 31 | 27267 – 27297 |
| **SWI – R2* subcortical** | Contains the iron content in subcortical structures | 14 | 25026 – 25039 |
| **T2 FLAIR WM hyperintensities** | Total volume of WM hyperintensities | 1 | 25781 |
| **Diffusion MRI FA (TBSS)** | Mean DTI fractional anisotropy (FA) in 48 ROIs | 48 | 25056 – 25103 |
| **Diffusion MRI MD (TBSS)** | Mean (MD) DTI mean diffusivity in 48 ROIs | 48 | 25104 – 25151 |
| **Diffusion MRI MO (TBSS)** | Mean DTI mode of anisotropy (MO) in 48 ROIs | 48 | 25152 – 25199 |
| **Diffusion MRI L1 (TBSS)** | Mean of the first DTI eigenvalue (L1) map in 48 ROIs | 48 | 25200 – 25247 |
| **Diffusion MRI L2 (TBSS)** | Mean of the second DTI eigenvalue (L2) map in 48 ROIs | 48 | 25248 – 25295 |
| **Diffusion MRI L3 (TBSS)** | Mean of the third DTI eigenvalue (L3) map in 48 ROIs | 48 | 25296 – 25343 |
| **Diffusion MRI ICVF (TBSS)** | Mean of NODDI intra-cellular volume fraction (ICVF) map in 48 ROIs | 48 | 25344 – 25391 |
| **Diffusion MRI OD (TBSS)** | Mean of NODDI orientation dispersion (OD) map in 48 ROIs | 48 | 25392 – 25439 |
| **Diffusion MRI ISOVF (TBSS)** | Mean of NODDI isotropic volume fraction (ISOVF) map in 48 ROIs | 48 | 25440 – 25487 |
| **Resting fMRI – nodes (D=25)** | Regional resting-state fMRI activity peak amplitude for coarse region definition (D=25) | 21 | 25754 |
| **Resting fMRI – nodes (D=100)** | Regional resting-state fMRI activity peak amplitude for fine region definition (D=100) | 55 | 25755 |
| **Full Resting fMRI FC – edges (D=25)** | Full correlation cortico-cortical resting-state functional connectivity for coarse region definition (D=25) | 210 | 25750 |
| **Partial Resting fMRI FC – edges (D=25)** | Partial correlation cortico-cortical resting-state functional connectivity for coarse region definition (D=25) | 210 | 25752 |
| **Task fMRI** | Contains the activation measures of an emotional task across regions extracted from group activation maps | 16 | 25040, 25042, 25044, 25046, 25048, 25050, 25052, 25054, 25761 – 25768 |

GM – grey matter; WM – white matter; CSF – cerebrospinal fluid; FA – fractional anisotropy; MD – mean diffusivity; MO – mode of anisotropy; ICVF – intra-cellular volume fraction; OD – orientation dispersion; ISOVF – isotropic volume fraction; TBSS – tract-based spatial statistics; RH – right hemisphere; LH – left hemisphere.

**Table S3.** List of confounds/covariates used in BWAS analysis.

| Confound/ Covariate | Confound/covariate Group Name | Confound/Covariate Name | UK Biobank Data Field | Original | Squared | Original (Gaussianised) | Squared (Gaussianised) | Total |
|---|---|---|---|---|---|---|---|---|
| Confound | Imaging measures | rfMRI head motion | 25741 | 1 | 1 | 1 | - | 3 |
| | | tfMRI head motion | 25742 | 1 | 1 | 1 | - | 3 |
| | | Head size scaling | 25000 | 1 | - | 1 | - | 2 |
| | | Bed position in scanner (x, y, z, and table) | 25756 - 25759 | 4 | 4 | 4 | 4 | 16 |
| | | Date-related drift | 53 | 10 | - | 10 | - | 20 |
| | | Imaging centre | 54 | 1 | - | - | - | 1 |
| Covariate | Demographics measures | Age | 34, 52, 53 | 1 | 1 | - | - | 2 |
| | | Sex | 31 | 1 | - | - | - | 1 |
| | | Age × Sex | - | 1 | - | - | - | 1 |
| | | Age squared × Sex | - | 1 | - | - | - | 1 |
| | Body measurements | Blood pressure (diastolic) | 4079 | 1 | 1 | - | - | 2 |
| | | Blood pressure (systolic) | 4080 | 1 | 1 | - | - | 2 |
| | | Height | 50 | 1 | 1 | - | - | 2 |
| | | Weight | 21002 | 1 | 1 | - | - | 2 |
| | Genetics information | Genetic matrix principal components | 22009 | 10 | - | - | - | 10 |
| | Lifestyle measures | Smoking status | 20116 | 1 | - | - | - | 1 |
| | | Alcohol consumption | 1568, 1578, 1588, 1598, 1608, 5364, 4407, 4418, 4429, 4440, 4451, 4462 | 1 | - | - | - | 1 |

**Table S4.** List of all significant IDPs associated with IPF risk genetic variants. 174 IDPs associated with the chromosome 17 (*17q21.31*) IPF variant and 19 IDPs associated with the chromosome (*DEPTOR gene*) IPF variant.

| Group IDP | IDP name | Chr. 17 | | Chr. 8 | |
|---|---|---|---|---|---|
| | | P-value | T-stat | P-value | T-stat |
| T1 Freesurfer cortical area | RH fusiform | 5.514e-24 | 10.109 | --- | --- |
| | LH fusiform | 9.738e-20 | 9.098 | --- | --- |
| | RH lingual | 4.685e-15 | 7.839 | --- | --- |
| | RH lateral occipital | 2.552e-14 | 7.623 | --- | --- |
| | LH lateral occipital | 7.138e-13 | 7.180 | --- | --- |
| | LH lingual | 3.358e-12 | 6.965 | --- | --- |
| | LH inferior parietal | 8.215e-11 | 6.499 | --- | --- |
| | RH cuneus | 6.639e-10 | 6.177 | --- | --- |
| | RH precuneus | 7.321e-09 | 5.785 | --- | --- |
| | RH inferior temporal | 2.357e-08 | 5.585 | --- | --- |
| | RH inferior parietal | 3.344e-08 | 5.524 | --- | --- |
| | LH supramarginal | 3.845e-08 | 5.499 | --- | --- |
| | LH rostral anterior cingulate | 4.671e-08 | 5.465 | --- | --- |
| | LH precuneus | 6.891e-08 | 5.395 | --- | --- |
| | RH post central | 6.953e-08 | 5.394 | --- | --- |
| | LH transverse temporal | 1.643e-07 | 5.237 | --- | --- |
| | RH posterior cingulate | 2.364e-07 | 5.169 | --- | --- |
| | RH supramarginal | 3.736e-07 | 5.083 | --- | --- |
| | LH post central | 6.842e-07 | 4.967 | --- | --- |
| | RH caudal anterior cingulate | 1.363e-06 | 4.831 | --- | --- |
| Diffusion MRI ICVF (TBSS) | Superior fronto-occipital fasciculus R | 6.279e-24 | 10.097 | --- | --- |
| | Posterior limb of internal capsule R | 7.745e-18 | 8.609 | --- | --- |
| | Superior fronto-occipital fasciculus L | 8.073e-18 | 8.604 | --- | --- |
| | Anterior corona radiata L | 9.289e-16 | 8.041 | --- | --- |
| | Anterior corona radiata R | 2.603e-15 | 7.913 | --- | --- |
| | Genu of corpus callosum | 2.879e-15 | 7.901 | 4.216e-09 | 5.877 |
| | Cingulum cingulate gyrus R | 4.098e-15 | 7.856 | --- | --- |
| | Posterior limb of internal capsule L | 1.280e-14 | 7.712 | --- | --- |
| | Anterior limb of internal capsule L | 4.856e-14 | 7.540 | --- | --- |
| | Cingulum cingulate gyrus L | 7.326e-14 | 7.486 | --- | --- |
| | Superior corona radiata R | 1.199e-13 | 7.421 | 9.847e-09 | 5.735 |
| | Superior longitudinal fasciculus R | 3.270e-13 | 7.286 | 3.813e-07 | 5.079 |
| | External capsule L | 9.074e-13 | 7.147 | --- | --- |
| | External capsule R | 1.865e-12 | 7.047 | --- | --- |
| | Superior corona radiata L | 6.795e-12 | 6.865 | 2.162e-07 | 5.186 |
| | Anterior limb of internal capsule R | 2.195e-11 | 6.695 | --- | --- |
| | Retrolenticular part of internal capsule L | 2.487e-11 | 6.677 | --- | --- |
| | Superior longitudinal fasciculus L | 1.425e-10 | 6.416 | 1.287e-06 | 4.843 |
| | Retrolenticular part of internal capsule R | 5.071e-10 | 6.219 | --- | --- |
| | Posterior corona radiata R | 5.111e-10 | 6.218 | --- | --- |
| | Body of corpus callosum | 5.549e-10 | 6.205 | 1.958e-11 | 6.712 |
| | Fornix cres+Stria terminalis R | 8.747e-10 | 6.133 | --- | --- |
| | Posterior corona radiata L | 3.126e-09 | 5.927 | --- | --- |
| | Fornix cres+Stria terminalis L | 7.065e-09 | 5.791 | --- | --- |
| | Cingulum hippocampus R | 1.493e-08 | 5.664 | --- | --- |
| | Splenium of corpus callosum | 1.172e-07 | 5.299 | 1.454e-07 | 5.260 |
| | Cingulum hippocampus L | 1.738e-07 | 5.227 | --- | --- |
| | Superior cerebellar peduncle R | --- | --- | 5.440e-08 | 5.438 |
| | Inferior cerebellar peduncle R | --- | --- | 7.066e-08 | 5.391 |
| | Cerebral peduncle L | --- | --- | 5.397e-07 | 5.013 |
| | Tapetum R | --- | --- | 9.379e-07 | 4.905 |
| | Posterior thalamic radiation R | --- | --- | 1.351e-06 | 4.833 |
| | Superior cerebellar peduncle L | --- | --- | 2.254e-06 | 4.730 |
| Diffusion MRI L3 (TBSS) | Anterior limb of internal capsule L | 2.619e-23 | 9.955 | --- | --- |
| | Anterior limb of internal capsule R | 1.863e-16 | 8.236 | --- | --- |
| | Genu of corpus callosum | 1.223e-12 | 7.106 | --- | --- |
| | Superior fronto-occipital fasciculus R | 1.377e-12 | 7.090 | --- | --- |
| | Superior corona radiata R | 9.901e-12 | 6.811 | --- | --- |
| | Superior fronto-occipital fasciculus L | 5.954e-11 | 6.548 | --- | --- |
| | Anterior corona radiata L | 4.719e-10 | 6.230 | --- | --- |
| | Anterior corona radiata R | 5.120e-08 | 5.449 | --- | --- |

| | | | | | |
|---|---|---|---|---|---|
| | Body of corpus callosum | 5.658e-08 | 5.431 | --- | --- |
| | Fornix cres+Stria terminalis L | 9.957e-08 | 5.329 | --- | --- |
| | Posterior limb of internal capsule R | 1.075e-07 | 5.315 | --- | --- |
| | External capsule L | 1.692e-07 | 5.232 | --- | --- |
| | Fornix | 4.171e-07 | 5.062 | --- | --- |
| | Posterior limb of internal capsule L | 1.388e-06 | 4.828 | --- | --- |
| | Splenium of corpus callosum | --- | --- | 2.941e-07 | 5.128 |
| Diffusion MRI MD (TBSS) | Anterior limb of internal capsule L | 1.346e-15 | 7.995 | --- | --- |
| | Genu of corpus callosum | 2.673e-15 | 7.910 | --- | --- |
| | Superior fronto-occipital fasciculus R | 9.956e-15 | 7.744 | --- | --- |
| | Anterior limb of internal capsule R | 1.035e-14 | 7.739 | --- | --- |
| | External capsule L | 9.229e-13 | 7.145 | --- | --- |
| | Anterior corona radiata L | 5.897e-12 | 6.885 | --- | --- |
| | Superior fronto-occipital fasciculus L | 1.688e-11 | 6.734 | --- | --- |
| | External capsule R | 1.747e-11 | 6.729 | --- | --- |
| | Posterior limb of internal capsule R | 4.889e-11 | 6.577 | --- | --- |
| | Superior corona radiata R | 1.614e-10 | 6.397 | 1.131e-06 | 4.868 |
| | Anterior corona radiata R | 2.258e-10 | 6.345 | --- | --- |
| | Superior longitudinal fasciculus R | 5.850e-10 | 6.197 | --- | --- |
| | Body of corpus callosum | 2.363e-09 | 5.973 | --- | --- |
| | Superior cerebellar peduncle L | 5.860e-09 | 5.823 | --- | --- |
| | Superior longitudinal fasciculus L | 1.123e-08 | 5.713 | --- | --- |
| | Superior corona radiata L | 1.842e-08 | 5.628 | --- | --- |
| | Cingulum cingulate gyrus L | 3.134e-08 | 5.535 | --- | --- |
| | Posterior limb of internal capsule L | 5.665e-08 | 5.431 | --- | --- |
| | Cingulum cingulate gyrus R | 1.178e-07 | 5.298 | --- | --- |
| | Fornix | 2.453e-07 | 5.163 | --- | --- |
| | Superior cerebellar peduncle R | 5.642e-07 | 5.004 | --- | --- |
| Diffusion MRI OD (TBSS) | Posterior corona radiata R | 3.308e-15 | 7.883 | --- | --- |
| | Superior longitudinal fasciculus L | 4.638e-14 | 7.546 | --- | --- |
| | Superior longitudinal fasciculus R | 3.847e-10 | 6.262 | --- | --- |
| | Posterior limb of internal capsule R | 5.561e-10 | 6.205 | --- | --- |
| | Posterior corona radiata L | 2.975e-09 | 5.935 | --- | --- |
| | Uncinate fasciculus R | 6.394e-09 | 5.808 | --- | --- |
| | Posterior thalamic radiation R | 3.489e-08 | 5.517 | --- | --- |
| | Cerebral peduncle R | 6.899e-08 | 5.395 | --- | --- |
| | Uncinate fasciculus L | 5.301e-07 | 5.016 | --- | --- |
| | Medial lemniscus R | 6.088e-07 | 4.990 | --- | --- |
| | Retrolenticular part of internal capsule R | 1.389e-06 | 4.828 | --- | --- |
| Diffusion MRI L1 (TBSS) | Superior longitudinal fasciculus L | 1.187e-14 | 7.722 | --- | --- |
| | External capsule R | 3.022e-14 | 7.601 | --- | --- |
| | Uncinate fasciculus R | 5.826e-14 | 7.516 | --- | --- |
| | Superior longitudinal fasciculus R | 1.322e-13 | 7.408 | --- | --- |
| | External capsule L | 3.390e-12 | 6.964 | --- | --- |
| | Anterior limb of internal capsule R | 1.022e-11 | 6.806 | --- | --- |
| | Posterior corona radiata R | 1.044e-11 | 6.803 | --- | --- |
| | Posterior limb of internal capsule R | 1.017e-10 | 6.467 | --- | --- |
| | Genu of corpus callosum | 1.168e-10 | 6.446 | --- | --- |
| | Superior fronto-occipital fasciculus R | 1.671e-10 | 6.391 | --- | --- |
| | Anterior corona radiata L | 2.493e-10 | 6.330 | --- | --- |
| | Uncinate fasciculus L | 3.916e-10 | 6.260 | --- | --- |
| | Anterior limb of internal capsule L | 9.649e-10 | 6.117 | --- | --- |
| | Superior corona radiata R | 8.302e-09 | 5.764 | --- | --- |
| | Superior corona radiata L | 2.687e-08 | 5.562 | --- | --- |
| | Fornix | 3.899e-08 | 5.497 | --- | --- |
| | Retrolenticular part of internal capsule L | 6.202e-08 | 5.414 | --- | --- |
| | Posterior corona radiata L | 7.373e-08 | 5.383 | --- | --- |
| | Anterior corona radiata R | 1.439e-07 | 5.262 | --- | --- |
| | Cerebral peduncle R | 2.013e-07 | 5.199 | --- | --- |
| | Medial lemniscus R | 3.327e-07 | 5.105 | --- | --- |
| | Cerebral peduncle L | 8.365e-07 | 4.928 | --- | --- |
| | Posterior thalamic radiation R | 1.660e-06 | 4.792 | --- | --- |
| | Medial lemniscus L | 2.197e-06 | 4.735 | --- | --- |
| SWI – R2* subcortical | T2star left putamen | 4.607e-14 | 7.547 | --- | --- |
| | T2star right putamen | 1.138e-12 | 7.116 | --- | --- |
| T1 global volumes | SIENAX white unnormalised volume | 3.449e-13 | 7.279 | --- | --- |
| | SIENAX brain unnormalised volume | 7.126e-13 | 7.180 | --- | --- |
| | SIENAX white normalised volume | 8.203e-13 | 7.161 | --- | --- |
| | SIENAX brain normalised volume | 4.734e-12 | 6.916 | --- | --- |
| | Anterior limb of internal capsule L | 1.583e-12 | 7.070 | --- | --- |

| | | | | | |
|---|---|---|---|---|---|
| Diffusion MRI MO (TBSS) | Superior longitudinal fasciculus L | 2.078e-08 | 5.607 | --- | --- |
| | Posterior limb of internal capsule R | 4.124e-08 | 5.487 | --- | --- |
| | Anterior limb of internal capsule R | 1.377e-07 | 5.270 | --- | --- |
| | Superior corona radiata R | 6.450e-07 | 4.978 | --- | --- |
| | Posterior corona radiata R | 7.930e-07 | 4.938 | --- | --- |
| T1 Freesurfer cortical thickness | LH inferior temporal | 3.541e-12 | 6.957 | --- | --- |
| | LH middle temporal | 4.976e-12 | 6.909 | --- | --- |
| | RH middle temporal | 4.298e-11 | 6.596 | --- | --- |
| | RH inferior temporal | 6.037e-09 | 5.818 | --- | --- |
| | LH inferior parietal | 9.340e-09 | 5.744 | --- | --- |
| | LH rostral anterior cingulate | 3.271e-08 | 5.528 | --- | --- |
| | LH pars triangularis | 3.606e-07 | 5.090 | --- | --- |
| | RH inferior parietal | 3.870e-07 | 5.076 | --- | --- |
| | RH lateral orbitofrontal | 4.145e-07 | 5.063 | --- | --- |
| | RH caudal anterior cingulate | --- | --- | 6.117e-07 | 4.989 |
| Diffusion MRI L2 (TBSS) | Genu of corpus callosum | 4.386e-12 | 6.927 | --- | --- |
| | External capsule L | 1.852e-10 | 6.376 | --- | --- |
| | Superior fronto-occipital fasciculus L | 4.289e-10 | 6.245 | --- | --- |
| | Anterior corona radiata L | 1.451e-08 | 5.669 | --- | --- |
| | External capsule R | 1.681e-08 | 5.644 | --- | --- |
| | Body of corpus callosum | 2.709e-08 | 5.561 | 5.874e-07 | 4.996 |
| | Anterior corona radiata R | 6.410e-09 | 5.808 | --- | --- |
| | Superior fronto-occipital fasciculus R | 1.760e-07 | 5.224 | --- | --- |
| | Fornix cres+Stria terminalis L | 3.532e-07 | 5.094 | --- | --- |
| | Superior cerebellar peduncle L | 8.359e-07 | 4.928 | --- | --- |
| | Fornix | 1.163e-06 | 4.863 | --- | --- |
| | Splenium of corpus callosum | --- | --- | 1.656e-07 | 5.236 |
| Diffusion MRI FA (TBSS) | Genu of corpus callosum | 6.834e-10 | 6.172 | --- | --- |
| | Anterior limb of internal capsule L | 1.998e-09 | 6.000 | --- | --- |
| | Superior fronto-occipital fasciculus L | 5.824e-09 | 5.824 | --- | --- |
| | Fornix cres+Stria terminalis L | 1.676e-07 | 5.233 | --- | --- |
| | Body of corpus callosum | 6.228e-07 | 4.985 | --- | --- |
| | Fornix | 8.635e-07 | 4.922 | --- | --- |
| | Splenium of corpus callosum | --- | --- | 6.346e-07 | 4.981 |
| T1 ROIs (FAST) | R intracalc cortex | 3.139e-09 | 5.926 | --- | --- |
| | L cerebellum crus II | 8.428e-09 | 5.761 | --- | --- |
| | R cerebellum crus II | 1.664e-08 | 5.645 | --- | --- |
| | L ventral striatum | 5.526e-08 | 5.435 | --- | --- |
| | R cerebellum VIIb | 9.999e-08 | 5.328 | --- | --- |
| | R planum polare | 1.561e-07 | 5.247 | --- | --- |
| | L cerebellum VIIb | 1.655e-07 | 5.236 | --- | --- |
| | L cerebellum VI | 2.012e-07 | 5.199 | --- | --- |
| | R cerebellum VIIIa | 2.196e-07 | 5.183 | --- | --- |
| | L cerebellum VIIIa | 8.386e-07 | 4.927 | --- | --- |
| T1 subcortical volumes (FIRST) | Left accumbens volume | 5.598e-08 | 5.433 | --- | --- |
| | Right hippocampus volume | 5.888e-07 | 4.996 | --- | --- |
| | Right pallidum volume | 9.705e-07 | 4.899 | --- | --- |
| | Left thalamus volume | 1.185e-06 | 4.859 | --- | --- |
| | Right thalamus volume | 1.339e-06 | 4.835 | --- | --- |
| Diffusion MRI ISOVF (TBSS) | Fornix | 1.342e-07 | 5.274 | --- | --- |
| | Superior cerebellar peduncle L | 1.857e-07 | 5.214 | --- | --- |
| | Superior cerebellar peduncle R | 1.926e-07 | 5.208 | --- | --- |
| | Posterior corona radiata L | 5.419e-07 | 5.012 | --- | --- |

RH – right hemisphere; LH – left hemisphere; R – right; and L – left.

**Table S5.** Co-localisation (COLOC) posterior probabilities (H0 – H4) for IDPs that were associated with chromosome 8 (*DEPTOR* gene) variant and chromosome 17 (*17q21.31*) variant in the BWAS model. The dominant hypothesis in each case is shown in bold. Seventeen of the nineteen IDPs that associated with the *DEPTOR* variant showed evidence of a likely shared causal variant with IPF (PP.H4 dominant). Two of these nineteen IDPs showed evidence of an association, but of no shared causal variant with IPF (one IDP with PP.H3 dominant and in the other IDP PP.H4 was dominant but PP.H4<0.8). For all the 174 IDPs associated with the chromosome 17 IPF variant, PP.H3 was the dominant (therefore a subset of 10 examples of these IDPs are shown).

| Chr. | Group IDP | IDP Name | PP.H0 | PP.H1 | PP.H2 | PP.H3 | PP.H4 |
|---|---|---|---|---|---|---|---|
| Chr. 8 | Diffusion MRI ICVF (TBSS) | Body of corpus callosum | 9.7e-15 | 2.3e-10 | 5.2e-06 | 0.122 | **0.878** |
| | | Genu of corpus callosum | 2.0e-12 | 4.7e-08 | 4.7e-06 | 0.110 | **0.890** |
| | | Superior corona radiata R | 7.6e-11 | 1.8e-06 | 7.0e-06 | 0.163 | **0.837** |
| | | Superior cerebellar peduncle R | 1.4e-09 | 3.3e-05 | 4.8e-06 | 0.112 | **0.888** |
| | | Superior cerebellar peduncle L | 3.4e-08 | 7.9e-04 | 4.7e-06 | 0.110 | **0.889** |
| | | Inferior cerebellar peduncle R | 1.1e-09 | 2.6e-05 | 4.2e-06 | 0.098 | **0.902** |
| | | Splenium of corpus callosum | 1.1e-09 | 2.5e-05 | 5.2e-06 | 0.121 | **0.879** |
| | | Superior corona radiata L | 7.1e-10 | 1.7e-05 | 1.4e-05 | 0.334 | 0.666 |
| | | Superior longitudinal fasciculus R | 2.7e-09 | 6.2e-05 | 4.9e-06 | 0.115 | **0.885** |
| | | Superior longitudinal fasciculus L | 4.5e-09 | 1.1e-04 | 4.8e-06 | 0.113 | **0.887** |
| | | Cerebral peduncle L | 3.5e-09 | 8.3e-05 | 4.6e-06 | 0.107 | **0.893** |
| | | Posterior thalamic radiation R | 1.5e-08 | 3.4e-04 | 5.3e-06 | 0.123 | **0.877** |
| | | Tapetum R | 3.4e-08 | 7.9e-04 | 3.9e-06 | 0.091 | **0.909** |
| | Diffusion MRI L2 (TBSS) | Splenium of corpus callosum | 6.0e-10 | 1.4e-05 | 5.9e-06 | 0.139 | **0.861** |
| | | Body of corpus callosum | 2.6e-09 | 6.0e-05 | 4.8e-06 | 0.111 | **0.888** |
| | Diffusion MRI L3 (TBSS) | Splenium of corpus callosum | 1.1e-09 | 2.7e-05 | 5.1e-06 | 0.119 | **0.881** |
| | T1 Freesurfer cortical thickness | Caudal anterior cingulate R | 1.2e-08 | 2.7e-04 | 5.3e-06 | 0.124 | **0.876** |
| | Diffusion MRI FA (TBSS) | Splenium of corpus callosum | 3.8e-09 | 8.8e-05 | 6.1e-06 | 0.142 | **0.858** |
| | Diffusion MRI MD (TBSS) | Superior corona radiata R | 7.7e-09 | 1.8e-04 | 2.2e-05 | **0.519** | 0.481 |
| Chr. 17 | T1 Freesurfer cortical area | Fusiform R | 8.1e-39 | 2.1e-27 | 2.7e-12 | **0.701** | 0.299 |
| | | Fusiform L | 1.8e-38 | 4.7e-27 | 2.6e-12 | **0.671** | 0.329 |
| | Diffusion MRI ICVF (TBSS) | Superior fronto-occipital fasciculus R | 3.6e-40 | 9.4e-29 | 2.5e-12 | **0.657** | 0.343 |
| | | Superior fronto-occipital fasciculus L | 1.8e-32 | 4.7e-21 | 2.7e-12 | **0.691** | 0.309 |
| | | Posterior limb of internal capsule R | 1.2e-34 | 3.2e-23 | 2.6e-12 | **0.663** | 0.337 |
| | | Anterior corona radiata L | 8.5e-31 | 2.2e-19 | 2.6e-12 | **0.667** | 0.333 |
| | | Anterior corona radiata R | 5.7e-30 | 1.5e-18 | 2.6e-12 | **0.666** | 0.334 |
| | Diffusion MRI L3 (TBSS) | Anterior limb of internal capsule L | 5.3e-39 | 1.4e-27 | 2.6e-12 | **0.678** | 0.322 |
| | | Anterior limb of internal capsule R | 2.1e-29 | 5.5e-18 | 2.9e-12 | **0.742** | 0.258 |
| | Diffusion MRI MD (TBSS) | Anterior limb of internal capsule L | 1.4e-27 | 3.6e-16 | 2.7e-12 | **0.688** | 0.312 |

**Table S6.** Mediation analysis coefficients, of *DEPTOR* variant to IDP association, through lung FVC. Results are shown for the 17 brain IDPs that were associated with the *DEPTOR* gene in BWAS and showed evidence of a likely shared causal variant with IPF through co-localisation. For each brain IDP, the first row represents the unstandardised regression coefficients and the standard error in parenthesis and the second row represents the t-stats, respectively for paths *a*, *b*, *c*, *c'*, and $a{\times}b$ (see Figure S2). The last column represents the result of the Sobel test (P-value for whether the indirect path $a{\times}b$ is different than zero).
($^{**}$) IDPs where significant partial mediation through FVC was found between *DEPTOR* gene and IDPs (Sobel test p<0.05 and $|c| > |c'|$).

| IDP name | *a* | *b* | *c'* (direct effect) | *c* (total effect) | *a×b* (indirect effect) | *P-value* (*a×b==0*) |
|---|---|---|---|---|---|---|
| FA – Splenium of corpus callosum | -0.0205 (0.0078) | 3.2x10$^{-5}$ (1.6x10$^{-4}$) | 8.63x10$^{-4}$ (1.9x10$^{-4}$) | 8.62x10$^{-4}$ (1.9x10$^{-4}$) | -6.5x10$^{-7}$ (1.6x10$^{-8}$) | --- |
| | -2.63 | 0.2 | 4.53 | 4.52 | -0.19 | |
| L2 – Body of corpus callosum | -0.0205 (0.0078) | -2.9x10$^{-6}$ (3.2x10$^{-7}$) | -1.46x10$^{-6}$ (3.9x10$^{-7}$) | -1.40x10$^{-6}$ (3.9x10$^{-7}$) | 6.0x10$^{-8}$ (2.4x10$^{-8}$) | --- |
| | -2.63 | -8.97 | -3.75 | -3.59 | 2.51 | |
| L2 – Splenium of corpus callosum | -0.0205 (0.0078) | -2.5x10$^{-7}$ (2.4x10$^{-7}$) | -1.445x10$^{-6}$ (2.9x10$^{-7}$) | -1.440x10$^{-6}$ (2.9x10$^{-7}$) | 5.2x10$^{-9}$ (5.7x10$^{-9}$) | --- |
| | -2.63 | -1.05 | -4.92 | -4.9 | 0.91 | |
| **L3 – Splenium of corpus callosum$^{**}$** | -0.0205 (0.0078) | 1.9x10$^{-6}$ (2.3x10$^{-7}$) | -1.06x10$^{-6}$ (2.8x10$^{-7}$) | -1.10x10$^{-6}$ (2.8x10$^{-7}$) | -3.9x10$^{-8}$ (1.6x10$^{-8}$) | 0.0129 |
| | -2.63 | 8.26 | -3.74 | -3.87 | -2.49 | |
| ICVF – Genu of corpus callosum | -0.0205 (0.0078) | 3.3x10$^{-4}$ (3.0x10$^{-4}$) | 2.00x10$^{-4}$ (3.6x10$^{-4}$) | 2.07x10$^{-4}$ (3.6x10$^{-4}$) | -6.7x10$^{-6}$ (7.0x10$^{-6}$) | --- |
| | -2.63 | 1.11 | 5.59 | 5.58 | -0.96 | |
| ICVF – Body of corpus callosum | -0.0205 (0.0078) | -6.3x10$^{-4}$ (2.5x10$^{-4}$) | 1.90x10$^{-3}$ (3.0x10$^{-4}$) | 1.89x10$^{-3}$ (3.0x10$^{-4}$) | 1.3x10$^{-5}$ (7.3x10$^{-6}$) | --- |
| | -2.63 | -2.56 | 6.42 | 6.46 | 1.76 | |
| **ICVF – Splenium of corpus callosum$^{**}$** | -0.0205 (0.0078) | -0.0020 (2.3x10$^{-4}$) | 1.26x10$^{-3}$ (2.8x10$^{-4}$) | 1.30x10$^{-3}$ (2.8x10$^{-4}$) | 4.1x10$^{-5}$ (1.6x10$^{-5}$) | 0.0128 |
| | -2.63 | -8.42 | 4.54 | 4.68 | 2.49 | |
| ICVF – Inferior cerebellar peduncle R | -0.0205 (0.0078) | 0.0027 (2.0x10$^{-4}$) | 1.40x10$^{-3}$ (2.47x10$^{-4}$) | 1.46x10$^{-3}$ (2.48x10$^{-4}$) | -5.5x10$^{-5}$ (2.2x10$^{-5}$) | --- |
| | -2.63 | 13.19 | 5.71 | 5.47 | -2.57 | |
| ICVF – Superior cerebellar peduncle R | -0.0205 (0.0078) | 0.0028 (1.7x10$^{-4}$) | 0.0010 (0.0002) | 9.9x10$^{-4}$ (0.0002) | -5.7x10$^{-5}$ (2.2x10$^{-5}$) | --- |
| | -2.63 | 16.72 | 5.24 | 4.93 | -2.59 | |
| ICVF – Superior cerebellar peduncle L | -0.0205 (0.0078) | 0.0027 (1.7x10$^{-4}$) | 0.0010 (0.0002) | 9.9x10$^{-4}$ (0.0002) | -5.5x10$^{-5}$ (2.1x10$^{-5}$) | --- |
| | -2.63 | 15.97 | 5.23 | 4.93 | -2.59 | |
| **ICVF – Cerebral peduncle L$^{**}$** | -0.0205 (0.0078) | -8.5x10$^{-4}$ (2.6x10$^{-4}$) | 1.58x10$^{-3}$ (0.0003) | 1.60x10$^{-3}$ (0.0003) | 1.8x10$^{-5}$ (8.7x10$^{-6}$) | 0.0425 |
| | -2.63 | -3.34 | 5.23 | 5.29 | 2.03 | |
| ICVF – Superior corona radiata R | -0.0205 (0.0078) | -6.6x10$^{-5}$ (2.6x10$^{-4}$) | 1.60x10$^{-3}$ (3.1x10$^{-4}$) | 1.601x10$^{-3}$ (3.1x10$^{-4}$) | -1.3x10$^{-6}$ (5.7x10$^{-6}$) | --- |
| | -2.63 | 0.25 | 5.27 | 5.27 | -0.24 | |
| **ICVF – Posterior thalamic radiation R$^{**}$** | -0.0205 (0.0078) | -0.0025 (0.0003) | 0.0020 (0.0004) | 0.0021 (0.0004) | 5.2x10$^{-5}$ (2.1x10$^{-5}$) | 0.0132 |
| | -2.63 | -8.02 | 5.26 | 5.39 | 2.48 | |
| ICVF – Superior longitudinal fasciculus R | -0.0205 (0.0078) | 1.8x10$^{-4}$ (2.8x10$^{-4}$) | 1.700x10$^{-3}$ (3.4x10$^{-4}$) | 1.704x10$^{-3}$ (3.4x10$^{-4}$) | -3.6x10$^{-6}$ (6.3x10$^{-6}$) | --- |
| | -2.63 | 0.63 | 5.14 | 5.13 | -0.58 | |
| ICVF – Superior longitudinal fasciculus L | -0.0205 (0.0078) | 2.5x10$^{-4}$ (2.8x10$^{-4}$) | 1.700x10$^{-3}$ (3.4x10$^{-4}$) | 1.705x10$^{-3}$ (3.4x10$^{-4}$) | -5.1x10$^{-6}$ (6.4x10$^{-6}$) | --- |
| | -2.63 | 0.90 | 5.13 | 5.11 | -0.80 | |
| **ICVF – Tapetum R$^{**}$** | -0.0205 (0.0078) | -0.0066 (4.8x10$^{-4}$) | 0.0023 (5.8x10$^{-4}$) | 0.0024 (5.8x10$^{-4}$) | 1.4x10$^{-4}$ (5.3x10$^{-5}$) | 0.0101 |
| | -2.63 | -13.85 | 3.98 | 4.20 | 2.57 | |
| Cortical thickness – caudal anterior cingulate RH | -0.0205 (0.0078) | -0.0158 (0.0026) | -0.0135 (0.0031) | -0.0131 (0.0031) | 3.2x10$^{-4}$ (1.4x10$^{-4}$) | --- |
| | -2.63 | -6.06 | -4.28 | -4.17 | 2.38 | |

#### Supplementary Figures

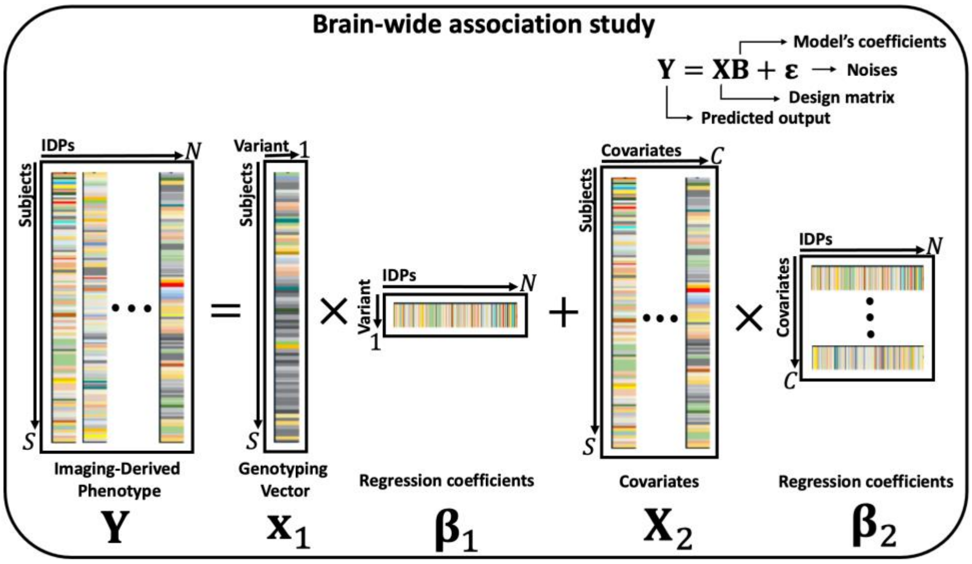


**Figure S1.** The general linear model used in the BWAS analysis. For each IPF variant, the IDPs matrix on the left-hand side, with dimensions Subjects × IDPs, is expressed as a linear combination of the genotyping vector for that variant and a covariates matrix. The analyses were run using PLINK[19].

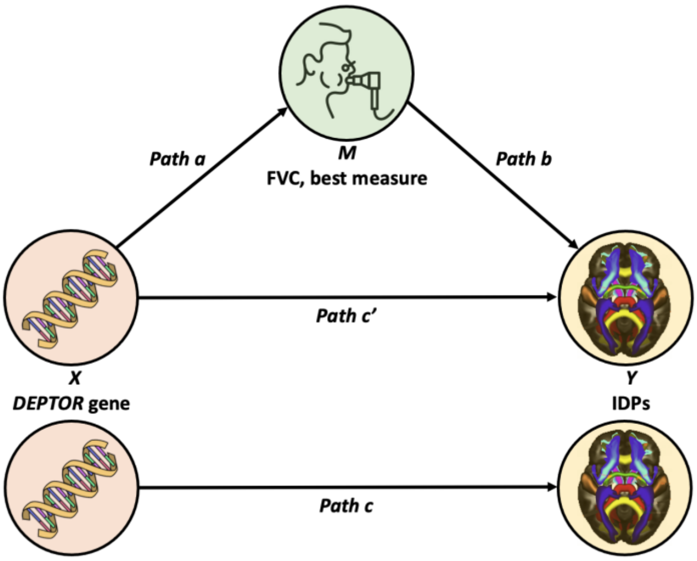


**Figure S2.** Schematic of the mediation model. Forced vital capacity (FVC) – best measure – is an intermediate, mediating variable $(M)$ between the *DEPTOR* gene $(X)$ and brain IDPs $(Y)$. In mediation analysis, the independent variable $(X)$ affects the mediator $(M)$, which in turn, affects the dependent variable $(Y)$. In other words, the relationship between the independent and dependent variables is assumed to be indirect. Path $a$ indicates the *DEPTOR* to FVC relationship and path $b$ FVC to brain IDPs relationships. The independent variable $X$ influences the dependent variable $Y$ directly (path $c'$) and indirectly (path $ab$) through a mediator $M$. The direct and indirect effects add to yield the total effect (path $c$) of $X$ on $Y$.

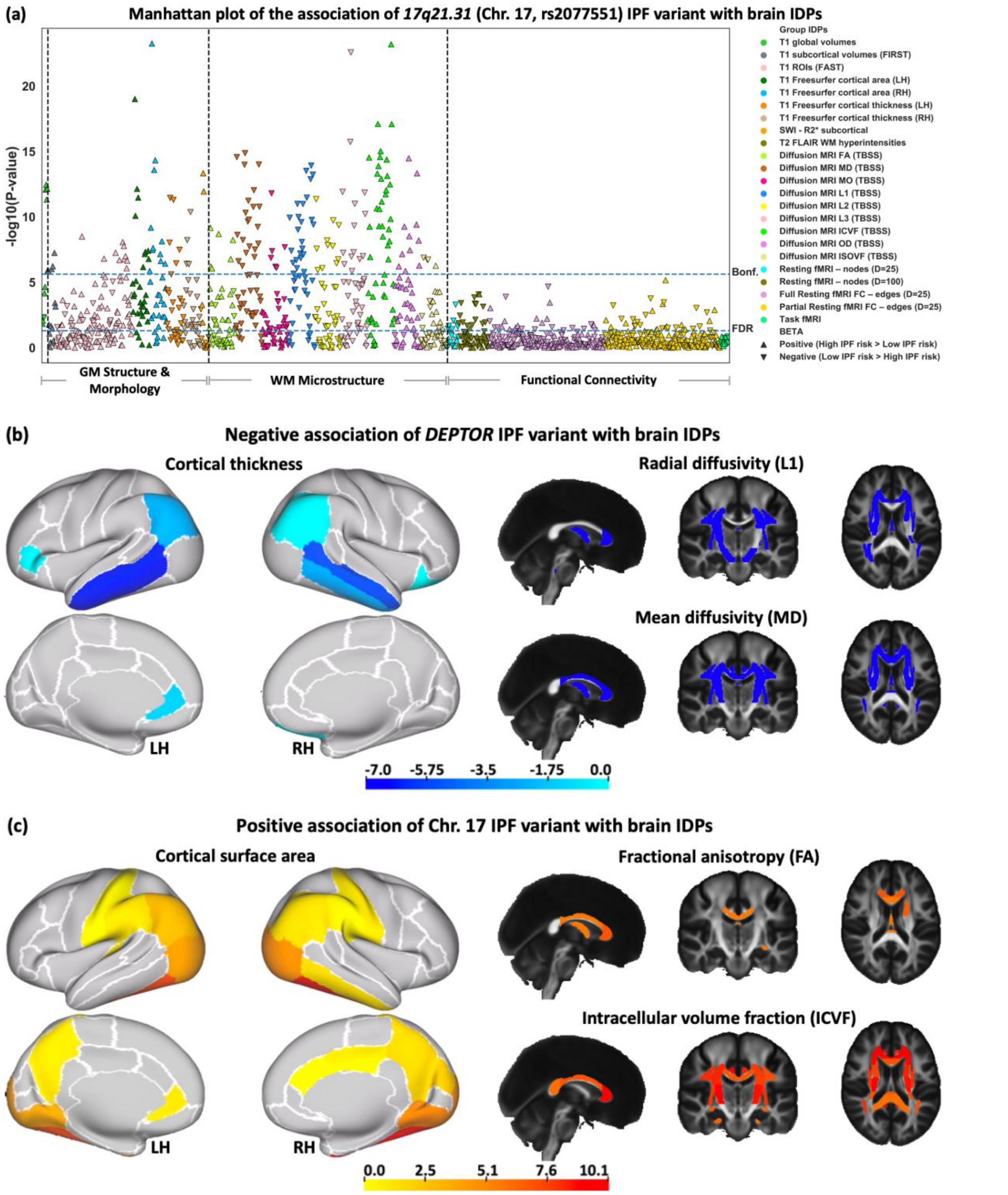


**Figure S3.** The association of chromosome 17 IPF variant with brain IDPs. **(a)** BWAS analysis of rs2077551 (*17q21.31* in chromosome 17). Points are coloured based on the group IDPs. The IDPs included in each group of features are available in Table S2. Triangles and inverted triangles are used to show the sign of the BETA in the BWAS. The dashed lines indicate the level of significance, i.e., FDR and Bonferroni thresholds (P-value<$2.36\times10^{-6}$). The vertical dashed lines separate the IDPs belongs to the grey matter (GM) structure and morphology, WM microstructure, and functional connectivity. A positive (negative) BETA means that a trait positively (negatively) associated with IPF risk. **(b)** Negative and **(c)** positive associations of neuroimaging features (IDPs) with IPF variant in rs2077551 (*17q21.31* in chromosome 17). T-stat maps of the associations are shown for the most significant IDPs (that survived Bonferroni correction) associated with the fibrosis variant. Positive t-stats correspond to positive association with high IPF risk. T1 Freesurfer-extracted cortical thickness, diffusion MRI L1, and diffusion MRI MD negatively associated with IPF variant in rs2077551 (*17q21.31* in chromosome 17). T1 Freesurfer-extracted cortical surface area, diffusion MRI FA, and diffusion MRI ICVF positively associated with the *17q21.31* IPF variant. In **(b-c)**, on the left, the white contours show the outline of the regions in Desikan-Killiany–Tourville (DKT) atlas and top and bottom panels show the lateral and medial view of the brain, respectively. In **(b-c)**, on the right, the standard HCP1065 FA image is used as background for the spatial maps (left is right). LH, Left hemisphere; RH, Right hemisphere.

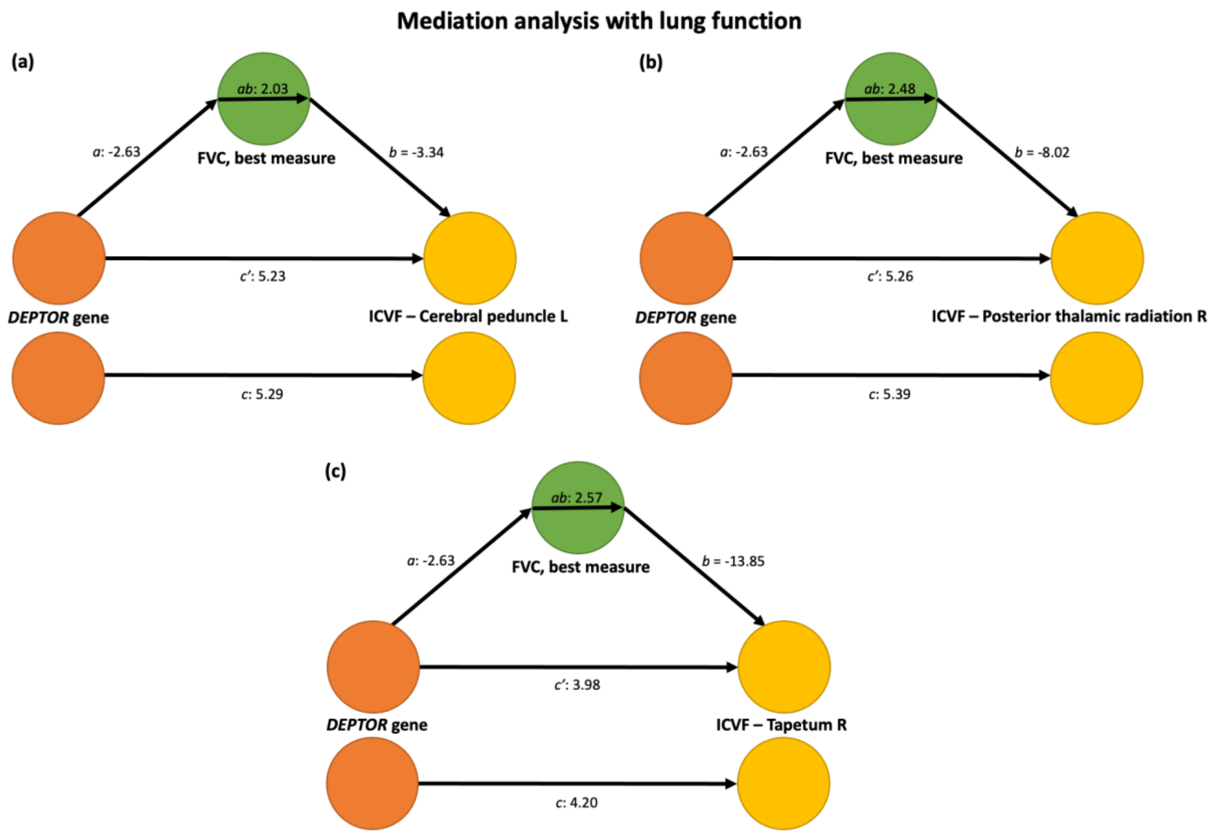


**Figure S4.** Summary of mediation analysis for the 3 (out of 5) IDPs with significant mediation through FVC, not shown in Main text. The path diagrams show a direct and indirect association between *DEPTOR* gene and the IDPs, a direct association between *DEPTOR* gene and FVC (best measure), and a direct path between FVC (best measure) and the IDPs. The IDPs are diffusion MRI **(a)** ICVF – Cerebral peduncle L, **(b)** ICVF – posterior thalamic radiation R, and **(c)** ICVF – tapetum R. The t-stats are shown for each path. All the path diagrams show a significant direct path between the *DEPTOR* gene and FVC (best measure), and a significant direct path between FVC and brain IDPs. All the direct and indirect effects between the *DEPTOR* gene and brain IDPs are significant. This means that FVC plays as a mediator to create an indirect association between the *DEPTOR* gene and brain IDPs.
